# COVCOG 2: Cognitive and Memory Deficits in Long COVID: A Second Publication from the COVID and Cognition Study

**DOI:** 10.1101/2021.10.27.21265563

**Authors:** Panyuan Guo, Alvaro Benito Ballesteros, Sabine P Yeung, Ruby Liu, Arka Saha, Lyn Curtis, Muzaffer Kaser, Mark P Haggard, Lucy G Cheke

## Abstract

Coronavirus disease 2019 (COVID-19), which is caused by severe acute respiratory syndrome coronavirus 2 (SARS-CoV-2), has been often characterized as a respiratory disease. However, it is increasingly being understood as an infection that impacts multiple systems, and many patients report neurological symptoms. Indeed, there is accumulating evidence for neural damage in some individuals, with recent studies suggesting loss of gray matter in multiple regions particularly in the left hemisphere. There are a number of mechanisms by which COVID-19 infection may lead to neurological symptoms and structural and functional changes in the brain, and cognitive problems are one of the most commonly reported symptoms in those suffering from Long COVID—the chronic illness following COVID-19 infection that affects between 10–25% of sufferers. However, there is as yet little research testing cognition in Long COVID. The COVID and Cognition Study is a cross-sectional/longitudinal study aiming to understand cognitive problems in Long COVID. The first paper from the study explored the characteristics of our sample of 181 individuals who had suffered COVID-19 infection, and 185 who had not, and the factors that predicted ongoing symptoms and self-reported cognitive deficits. In this second paper from the study, we assess this sample on tests of memory, language and executive function. We hypothesize that performance on “objective” cognitive tests will reflect self-reported cognitive symptoms. We further hypothesize that some symptom profiles may be more predictive of cognitive performance than others, perhaps giving some information as to mechanism. We found a consistent pattern of memory deficits in those that had suffered COVID-19 infection, with deficit increasing with severity of self-reported ongoing symptoms. Fatigue/Systemic symptoms during the initial illness and ongoing neurological symptoms were predictive of cognitive performance.

## 1 Introduction

COVID-19 has traditionally been considered a respiratory disease. However, around 35% of patients—and up to 85% of those who become severely ill—report neurological symptoms including headache, dizziness, myalgia or loss of taste and smell (e.g., Mao et al. 2020). The most well known neurological symptom—alteration in taste or smell (anosmia/dysgeusia)—is also one of the most common symptoms of the disease (e.g., Lechien et al. 2020), often the first symptom to manifest (Mao et al. 2020; Romero-Sánchez et al. 2020) and last to abate (Lechien et al. 2020).

There is accumulating evidence that COVID-19 is associated with neural damage, particularly in the presence of neurological symptoms (Helms et al. 2020; Kandemirli et al. 2020). Post-mortem studies of patients who have died of COVID-19 show evidence for ischemic lesions, and indications of neuro-inflammation (Matschke et al. 2020). Multiple studies have indicated abnormalities such as hemorrhagic lesions in the orbitofrontal cortex (Le Guennec et al. 2020), medial temporal lobe and hippocampus (Moriguchi et al. 2020; Poyiadji et al. 2020), bilateral thalami, and subinsular regions (Poyiadji et al. 2020). The changes may be functional as well as structural, with nearly 90% of electroencephalography (EEG) studies conducted in COVID-19 patients revealed epileptiform discharges, mostly within the frontal lobes (Galanopoulou et al. 2020). A study using the UK Biobank cohort conducted structural and functional brain scans before and after infection with COVID-19 on 394 patients compared with 388 matched controls who had not experienced COVID-19 infection (Douaud et al. 2021). Significant loss of gray matter was identified in the parahippocampal gyrus, lateral orbitofrontal cortex and insula, and was notably concentrated in the left hemisphere. An analysis of the small subset of this sample (*N* = 15) who had been hospitalized indicated more severe gray matter loss in these participants, particularly in the left cingulate cortex, and right amygdala and hippocampus. Bougakov, Podell, and Goldberg (2021) have argued that depending on the mechanism and location of neural damage, there are a number of cognitive deficits that might be expected to be detectable in COVID-19 patients. For example, SARS-CoV-2 may be able to attack the brain directly perhaps via the olfactory nerve (Lechien et al. 2020; Politi, Salsano, and Grimaldi 2020) causing encephalitis. On the other hand, severe hypoxia from respiratory issues may induce hypoxic/anoxic encephalopathy (Guo et al. 2020). The unusual clotting seen in COVID-19 may be associated with acute ischemic and hemorrhagic cerebrovascular events (CVAs: Beyrouti et al. 2020; Kubánková et al. 2021; Li et al. 2020; Wang et al. 2020) leading to more lasting brain lesions. Finally, a maladaptive immune response to infection can negatively impact neural systems via hemorrhagic encephalopathy (Das, Mukherjee, and Ghosh 2020; Poyiadji et al. 2020) or peripheral neuropathy (e.g., Guillain-Barre syndrome; Alberti et al. 2020; Whittaker, Anson, and Harky 2020; Zhao et al. 2020).

Much of the evidence suggesting that cognitive dysfunction may occur following COVID-19 infection comes from those who experience “post-COVID-19 syndrome”/ “post-acute sequalae SARS CoV-2” (PASC) / “Long COVID”. The National Institute for Health and Care Excellence (NICE) guidelines describe “post-COVID-19 syndrome” as “*Signs or symptoms that develop during or after infection consistent with COVID-19, continue for more than 12 weeks and are not explained by an alternative diagnosis”* (NICE 2020). Disagreements exist as to the exact symptoms, longevity and severity required to qualify for a diagnosis of PASC, making it difficult to ascertain prevalence precisely. However, estimates range from 10–25% of COVID-19 sufferers going on to having some degree of chronic illness (e.g.,Cirulli et al. 2020; Ding et al. 2020; Nehme et al. 2021; ONS 2021; Sudre et al. 2020; Tenforde et al. 2020). The patient-created term “Long COVID” has increasingly been used as an umbrella term to describe this highly heterogenous condition (Callard and Perego 2021).

Cognitive dysfunction is one of the most common symptoms reported in research into Long COVID, occurring in around 70% of patients (Bliddal et al. 2021; Cirulli et al. 2020; Davis et al. 2021; Ziauddeen et al. 2021) in many cases appearing second only to fatigue. In one study, 86% of participants indicated that cognitive dysfunction and/or memory impairment was impacting their ability to work (Davis et al. 2021). In our first paper in the COVID and Cognition study (Guo et al. 2021) we found a similar prevalence of cognitive symptoms to previous studies, with 77.8% reporting difficulty concentrating, 69% reporting brain fog, 67.5% reporting forgetfulness, 59.5% reporting tip-of-the-tongue (ToT) word-finding problems and 43.7% reported semantic disfluency (saying or typing the wrong word). In that analysis, we found that experience of chronic fatigue-like (“Fatigue/Systemic”) and neurological symptoms during the first 3 weeks significantly predicted experience of cognitive symptoms later in the subsequent illness. Those individuals experiencing ongoing “Cardio-Pulmonary”, “Neurological” and “Gastrointestinal/Autoimmune” symptoms were also found to be more likely to be experiencing cognitive symptoms.

It is often difficult to ascertain to what extent quite broadly defined self-reported cognitive deficits such as “difficulty concentrating” and “brain fog” translate into measurable changes in cognitive performance. While there are multiple lines of evidence to suggest that individuals suffering from Long COVID experience cognitive symptoms, there is—to date— little research objectively measuring cognition post-COVID-19.

Alemanno and colleagues (2021) investigated cognitive function in the post-acute phase (1 month after discharge) in COVID-19 patients that had suffered from severe illness. Using the Montreal Cognitive Assessment (MoCA) they showed that 80% of patients showed indications of cognitive deficit, particularly in memory, executive function and language. Similarly, Helms and colleagues (2020) found that, at discharge from intensive care unit (ICU), 33% of patients showed evidence of dysexecutive syndrome, with symptoms such as inattention, disorientation, or poorly organised movements in response to command. In their study of 29 patients (average age 65) presenting at least one new neurological symptom since COVID-19 infection, Hosp and colleagues (2021) found that cognitive performance may be linked to neurological abnormalities and symptoms. Positron emission tomography (PET) analysis revealed predominant frontoparietal hypometabolism, correlating to lower scores on the MoCA and extended neuropsychological testing. In particular, COVID-19 patients showed deficits in tests of verbal memory and executive functions. One issue with all of these studies data is limited to severely ill patients, mostly of older age (65+). It is thus difficult to determine whether these deficits are specific to COVID-19, or a more general response to acute respiratory distress (ARD) and ventilation. It is known, for example, that survivors of critical illness are known to suffer long-term cognitive impairment (Ehlenbach et al. 2010; Hopkins et al. 1999; Iwashyna et al. 2010; Jackson et al. 2003; Pandharipande et al. 2013) particularly if they experience delirium (e.g., Girard et al. 2010; Pandharipande et al. 2013). Thus it is important to establish to what extent cognitive dysfunction is a feature of post-COVID-19 pathology, or merely reflective of the large number of COVID-19 patients that experience ARD. Furthermore, it must be investigated whether these deficits extend into younger populations. In an early indication that this might be the case, Almeria and colleagues (2020) assessed younger (aged 24–60) patients 10–40 days post discharge, of which only 20% had been in intensive care, but 60% required oxygen. They found that those reporting neurological symptoms had lower performance on attention, memory and executive function, once again suggesting a degree of association between symptomatology and degree of cognitive deficit.

In a very large study using 81,337 participants in the Great British Intelligence Test (GBIT; mean age 46.75), Hampshire and colleagues (2021) compared participants who reported having had COVID-19 infection to concurrently tested control participants. The authors were able to conduct an analysis of association between symptom severity and cognitive performance controlled for age, gender, education level, income, racial-ethnic group, and pre-existing medical disorders. Of 12,689 participants that suspected that they had had COVID-19, 326 had a positive test, and 192 were hospitalized. Participants who had received a positive test had a lower global score and this deficit scaled with severity of initial respiratory illness: There was a substantial effect size for people who had been hospitalized, but also a clear effect for mild but biologically confirmed cases who reported no breathing difficulties. The largest effect sizes were seen in tests of verbal reasoning, multi-stage planning and spatial attention. Most participants had fully recovered at the time they took the test, however 24% of those with test-confirmed COVID-19 reported residual symptoms. Controlling for residual symptoms, respiratory severity during the initial illness remained a strong predictor of global cognitive performance, while presence of ongoing symptoms did not predict significant variance. There was no significant association between time since illness and cognitive performance, however this analysis excluded those with ongoing symptoms.

Graham and colleagues (2021) investigated cognition and quality of life measures in 100 non-hospitalized patients (mean age 43) presenting to a neuro-COVID clinic with neurological symptoms persisting for at least 6 weeks from symptom onset. These patients reported a median of five neurologic symptoms and over 80% reported having experienced brain fog. Some, but not all, of these symptoms had resolved at the time of cognitive assessment. A subset of participants were assessed on the National Institutes of Health (NIH) Toolbox covering processing speed, attention and executive memory, executive function and working memory, and compared to established baselines. The authors reported 53% of participants as having abnormal findings, with short-term memory and attention being most commonly impaired. Participants also had significantly reduced cognition- and fatigue-related quality of life indices. However, given that performance in this study was compared to established baselines rather than a control group, it is difficult to be confident of the proportion of the seen deficit that is attributable to COVID-19 rather than the general stress and disruption caused by the pandemic.

Despite it being probable that there is a relationship between COVID-19 infection, neurological symptoms and cognitive dysfunction, many questions remain about the specific nature of the cognitive impairment in Long COVID. We distinguish three main ones that drive our research programme and which it attempts to answer: First, what are the associations between reported symptoms and cognitive outcomes? Second, given the heterogenous nature of Long COVID, is that diversity reflected in a diversity of cognitive issues, or is there a specific sub-phenotype of Long COVID that is associated with cognitive deficits? Finally, are those that report “subjective” cognition and memory complaints more likely to demonstrate impairments in “objective” cognitive assessments of the same functions?

Here we report on the first stage of a mixed cross-sectional/longitudinal study—The COVID and Cognition Study (COVCOG)—aimed at understanding cognition following COVID-19 infection relative to that of concurrently tested controls. Using the online assessment platform Gorilla (www.gorilla.sc), we set out to bring together information about symptom profiles both during and following acute infection and detailed analysis of cognitive performance across a range of domains including memory, language and executive function. The aims of this study do not include identifying a specific mechanism of cognitive deficit (as that requires types of tests and analysis not feasible in an online study) but rather to “map the terrain”, providing sufficient breadth and detail of mechanism-relevant information to facilitate and inform future mechanistic investigation.

The first aim of this investigation was to ascertain whether differences could be found in cognitive performance between those that had and had not suffered COVID-19 infection. Problems with memory and with speech and language are the most commonly reported cognitive symptoms (after “brain fog”) in Long COVID—affecting around 70% and 40% of sufferers respectively (Davis et al. 2021). Given this, we hypothesize that where cognitive differences exist, these will be larger, or more likely, in tests assessing memory or language relative to those assessing (for example) executive function.

A second hypothesis, following previous findings (e.g., Hampshire et al. 2021; Hosp et al. 2021) is that degree of cognitive deficit will relate to severity and nature of initial illness. In particular, it seems likely that number and severity of neurological symptoms during the initial illness may be indicative of the degree of impact of the disease on neural function (whether that be via direct infection, inflammation or CVA or another route), which would be mostly likely to result in subsequent cognitive deficits. Our previous publication on this study (Guo et al. 2021) found that ongoing cognitive symptoms were predicted by Fatigue/Systemic, Neurological and Respiratory/Infectious (e.g., cough, fever, loss of taste and smell) symptoms experienced during the acute illness. We predict that similar associations will be found between symptom factors during the initial illness and performance on cognitive tasks, and that these may be most pronounced for neurological symptoms.

We further hypothesize that not just the presence but the nature of ongoing illness will be associated with cognitive deficits. We predict that those with severe ongoing symptoms will be more likely to show concomitantly more severe deficits in cognitive tasks. Our first paper from this study found that ongoing Cardio-Pulmonary, Neurological and “Gastrointestinal/Autoimmune” symptoms were associated with greater cognitive symptoms. We hypothesize that these symptom factors will be similarly associated with performance on cognitive tests.

Finally, we predict that any deficits will be greatest in those individuals experiencing ongoing cognitive symptoms. Indeed, we might expect those reporting specific cognitive symptoms (e.g., “forgetfulness”) to be particularly impaired on tests of cognition that assess the associated skill (e.g., memory).

## 2 Methods

### 2.1 Participants

A total of 421 participants aged 18 and over were recruited through word of mouth, student societies and online/social media platforms such as the Facebook *Long COVID Support Group* and the *Prolific* recruitment site. Of these 181 (130 female) had experienced COVID-19 infection (65 test-confirmed, 96 suspected) and 185 (118 female) had not. A further 55 had “unknown” infection status (did not think they had had COVID-19, but had had an illness that could potentially have been). Of those that had had COVID-19, 42 (29 female) had recovered by the time of test (“Recovered group”, *R*), 53 (36 female) continued to experience mild or moderate ongoing symptoms (“Ongoing (Mild/<u>M</u>oderate) group”, *C+*), and 66 (54 female) experienced severe ongoing symptoms (“Ongoing (Severe) group”, *C++*). A Further 20 participants were too early in the illness to indicate ongoing symptoms. Full details of our sample—including demographic and medical history characterisations—are provided in our previous publication on this study (Guo et al. 2021).

### 2.2 Procedure

The study received favorable ethical opinion from University of Cambridge Department of Psychology Ethics Committee (PRE.2020.106, 8/9/2020). This is a mixed cross-sectional/longitudinal online study conducted using Gorilla (www.gorilla.sc). The results reported here are for the baseline session of the study only. The baseline session consisted of a questionnaire covering demographics, previous health and experience of COVID-19, followed by a series of cognitive tests.

Participants answered questions relating to their age, sex, education level, country of permanent residence, ethnicity, and profession. They were then asked a series of questions relating to their medical history, and health-related behaviours (such as smoking and exercise). Next they were asked for details of their experience of COVID-19. COVID-status was established based on their response to a series of questions (starting with “Have you had COVID-19?”) and their response to a series of questions regarding presence and severity of ongoing symptoms. Full details of the questionnaires and grouping-dynamics are provided in our previous publication on this study (Guo et al. 2021). Finally, participants were asked to give details on a large number of individual symptoms during three time periods: the initial 3 weeks, “in the time since then”, and the past 1–2 days. Participants were also asked to report 5-point Likert scale, from very bad (1) to very good (5) on how current symptom severity was on the day of test.

### 2.3 Cognitive Tests

Figure 1 shows the 6 cognitive tasks that were presented. All participants completed tasks a–d and f, while only the “No COVID” group completed task e.

**Figure 1.**
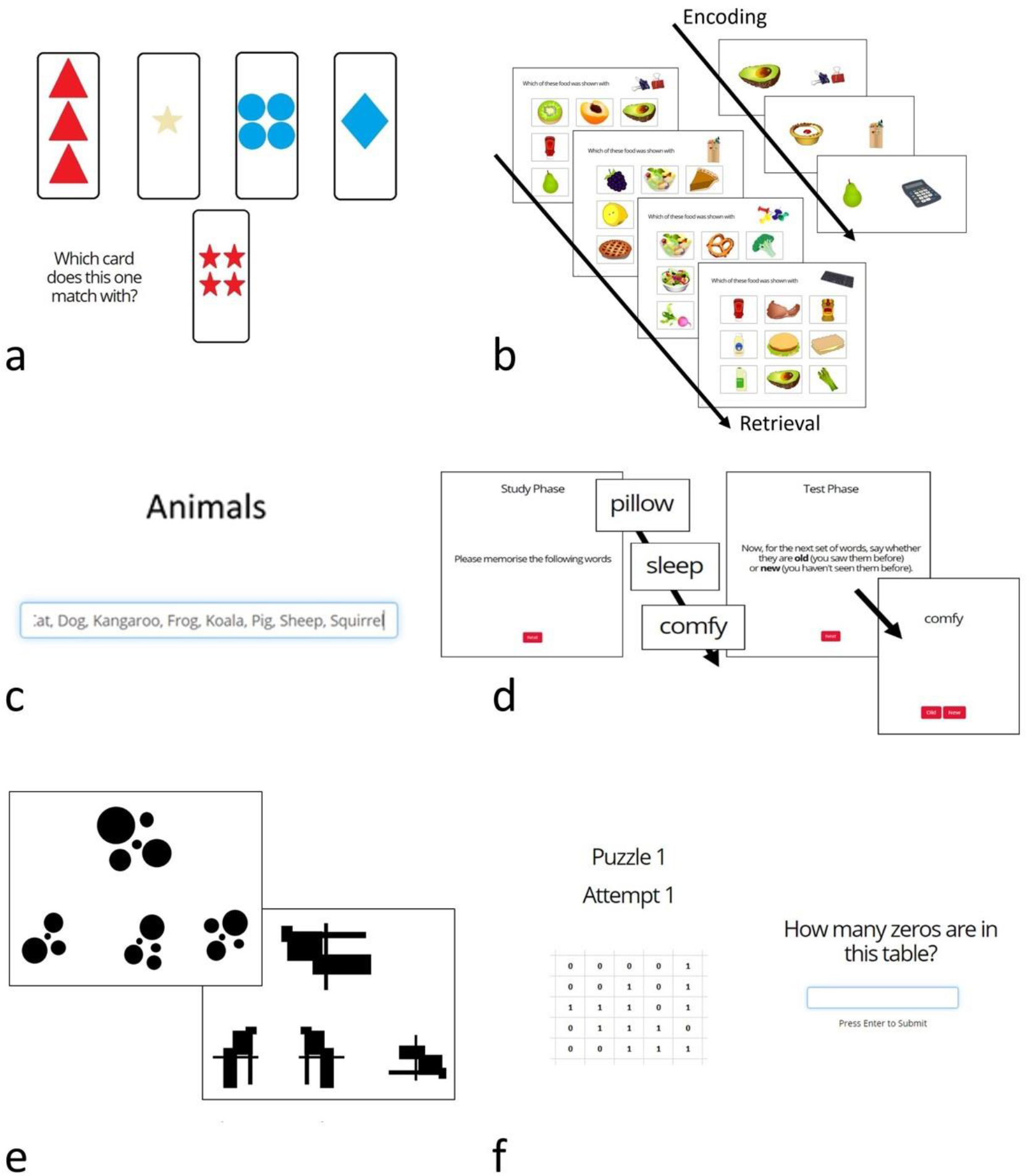
Cognitive Tasks. a) Wisconsin Card Sorting Test; b) Pictorial Associative Memory Test; c) Category Fluency Test; d) Word List Recognition Memory Test; e) 2D Mental Rotation Test; f) Number Counting Test (Attention/Bot Check).

#### 2.3.1 Word List Recognition Memory Test (Figure 1d)

Participants were shown a list of 16 words one by one with the instruction to memorize as many as possible. They were then shown 32 (16 old, 16 new) words and asked to report which had been on the original list. Target and distractor words were scored and matched for imagery and concreteness. The dependant variables on this task were % correct, d’ and reaction time (RT).

#### 2.3.2 Pictorial Associative Memory Test (Figure 1b)

Participants were required to memorize a series of 17 stationery and food-item pairs each displayed on the screen for three seconds. The recall phase took place immediately thereafter and involved 15 trials, each of which presented an item of stationery and asked participants to select the associated food-item from 9 options. The dependant variables were % correct and reaction time.

#### 2.3.3 Category Fluency Test (Figure 1c)

Participants were presented with the category word “Animals” and had 1 minute to type every example of that category they could think of. The words were entered into a scrolling text box such that, after around 6 words, earlier words started to move out of view. Dependant variables were number of correct words, % produced words that were correct, number of incorrect (unrelated) words (e.g., “table”), number of incorrect (related) words (e.g., “fur”), and number of repetitions.

#### 2.3.4 2D Mental Rotation Test (Figure 1e)

Participants were presented with 16 trials in which they saw an abstract image and had to select which of three possible options represented that image rotated. This is a test of visual working memory. Outcome variables were % correct and reaction time.

#### 2.3.5 Wisconsin Card Sorting Test (WCST) (Figure 1a)

This executive function (EF) task assesses task switching and inhibition. Across 64 trials, participants are required to match a given card to one of four cards on the basis of either colour, shape or number. They are not explicitly told the matching rule, but must infer this from the feedback on their choices. Every few trials the rule changes and participants must find and follow the new rule based on feedback.

#### 2.3.6 Number Counting Test (Figure 1f)

This task was included in the baseline as an attention / “bot” control for data quality. It presents a grid of 1s and 0s and asks the participants to count the 0s. This is not cognitively difficult but requires concentration. Because the grid is an image, this is also difficult for most AIs. Participants are given 3 attempts at this task. The numbers given by participants giving 3 incorrect answers were manually checked. If the numbers appeared to be genuine attempts (i.e., close but incorrect) then the participant was considered genuine and included in the dataset. No participants were removed due to failing this task.

#### 2.3.7 Relational Reasoning Test

Across 35 trials participants were shown a 3×3 matrix of images with one missing, and asked to select from 4 options which image should fill the gap. This task was given only to the No COVID group and was intended as a means by which to IQ-match control participants for potential pre-post infection longitudinal explorations. Data from this task are not reported in this paper.

### 2.4. Data processing and Analysis

Analyses were conducted using IBM SPSS Statistics for Windows, Version 23.0. We describe quantitative variables using means and standard deviations, and numbers and percentages for qualitative variables. Sidak’s correction for multiple comparisons was employed where appropriate, and both corrected and uncorrected analyses are shown.

As there were a large number of cognitive test variables, we reduced these via factor analysis to produce 4 factors representing Executive Functions (Performance), Executive Functions (Reaction Time), Memory and Category Fluency. Analyses were conducted first on these factors, to give an overview of pattern of cognitive performance, and then on the individual variables to give a more detailed picture.

We investigated differences in cognitive performance first dividing the sample into two groups (COVID/No COVID), second subdividing the COVID group by symptom longevity and severity (Recovered, Ongoing mild infection, and Ongoing severe infection). Where parametric analysis was not appropriate, we employed the Pearson’s chi-square (*χ^2^*) for categorical variables and the Mann-Whitney and Kruskal-Wallis test for continuous variables depending on the number of COVID groups. To explore what variables were associated with infection or ongoing symptoms we employed various independent multinomial logistic regression models (backward method). To investigate differences between groups (COVID/No COVID; Recovered / Ongoing mild / Ongoing severe) and the outcome of the cognitive tasks we employed independent t-test/Mann-Whitney and ANOVA/Kruskal-Wallis. We also performed general linear models controlling for sex, age, country, and education level. We also examined whether any total score from the cognitive tasks could be associated with variance in initial illness severity (asymptomatic/very mild, mild, and moderate/severe) using independent simple regression models.

As reviewed in detail in our previous publication (Guo et al. 2021), we used exploratory principal component analysis to cluster the symptoms experienced during the acute infection, and the symptoms subsequently experienced since that time. We identified 5 factors for symptoms experienced during the first 3 weeks of illness. These included a “Fatigue/Systemic” factor which was characterized by fatigue, chest pain/tightness and muscle/body pains; a “Gastrointestinal” factor, characterized by diarrhea, nausea and vomiting, a “Respiratory/Infectious” factor characterized by fever, cough, and breathing issues and a “Dermatological” factor characterized by rash, itchy welts, and foot sores. For symptoms experienced in the time since the initial illness, 6 factors were identified: A “Neurological” factor characterized by disorientation, confusion and delirium, a “Gastrointestinal/Autoimmune” factor characterized by hot flushes, nausea and diarrhea, a “Cardio-Pulmonary” factor characterized by breathing issues, chest pain/tightness and fatigue, a “Dermatological/Fever” factor characterized by face/lips swelling, foot sores, and itchy welts, an “Appetite Loss” factor characterized by weight loss and loss of appetite, and finally, a “Mood” factor characterized by depression, anxiety and vivid dreams. To assess currently experienced symptom factors, we employed the *sum scores by factor* method using the “since then” symptom factors as a base. We used linear multiple regression models (backward method) to test whether ongoing factors predicted performance on cognitive tests.

## 3 Results

### 3.1 Factor Analysis of Cognitive Variables

The cognitive task variables were a priori divided into two groups: language and memory (incorporating all Word List, Associative Memory and Category Fluency variables), and executive functions (including all WCST and 2D Mental Rotation variables), and factor analyses were conducted on these separately. Each exploratory factor analysis (EFA) was limited to two factors. Two items (one in each analysis: WCST perseverative error reaction time and Category Fluency repetitions) which did not load into any factor and were removed. The re-run analyses explained 48.9% and 58.9% of variance respectively. We thus ended with four performance factors: Executive Functions Performance (including score and errors for WCST and performance on 2D Mental Rotation), Executive Functions Reaction Times (including all reaction times from both EF tasks), Memory (including all variables from both Word List and Associative Memory) and Category Fluency (including all Category Fluency variables). See supplementary Table 1 for rotated component matrix.

Ten extreme outliers (identified by Q plot) were removed from each of the Category Fluency and EF RT factors to bring skewness and kurtosis within acceptable bounds (Category: skew = −0.623 (0.139); kurtosis = −0.181 (0.276); EF RT: skew = −0.508 (0.138); kurtosis = −0.153 (0.274)). Similarly, 9 extreme outliers (identified by Q plot) were removed from the Memory factor (skew = −0.623 (0.139); kurtosis = −0.181 (0.276)).

### 3.2 COVID-19 and Cognition

#### 3.2.1 Memory and Word Finding

A first analysis was run using the task factors comparing the “COVID” and “No COVID” groups, there was a significant negative influence of COVID-19 infection on memory performance, even when controlling for age, sex, country and education level (*F*(1,304) = 10.903, *p* = .001).

There was also a significant difference between groups on the Category Fluency factor (*F*(1,307) = 6.297, *p* = .013, *ή_p_^2^* = .02) but this disappeared when controlling for demographic variables (see Table 1, Figure 2).

**Table 1.**
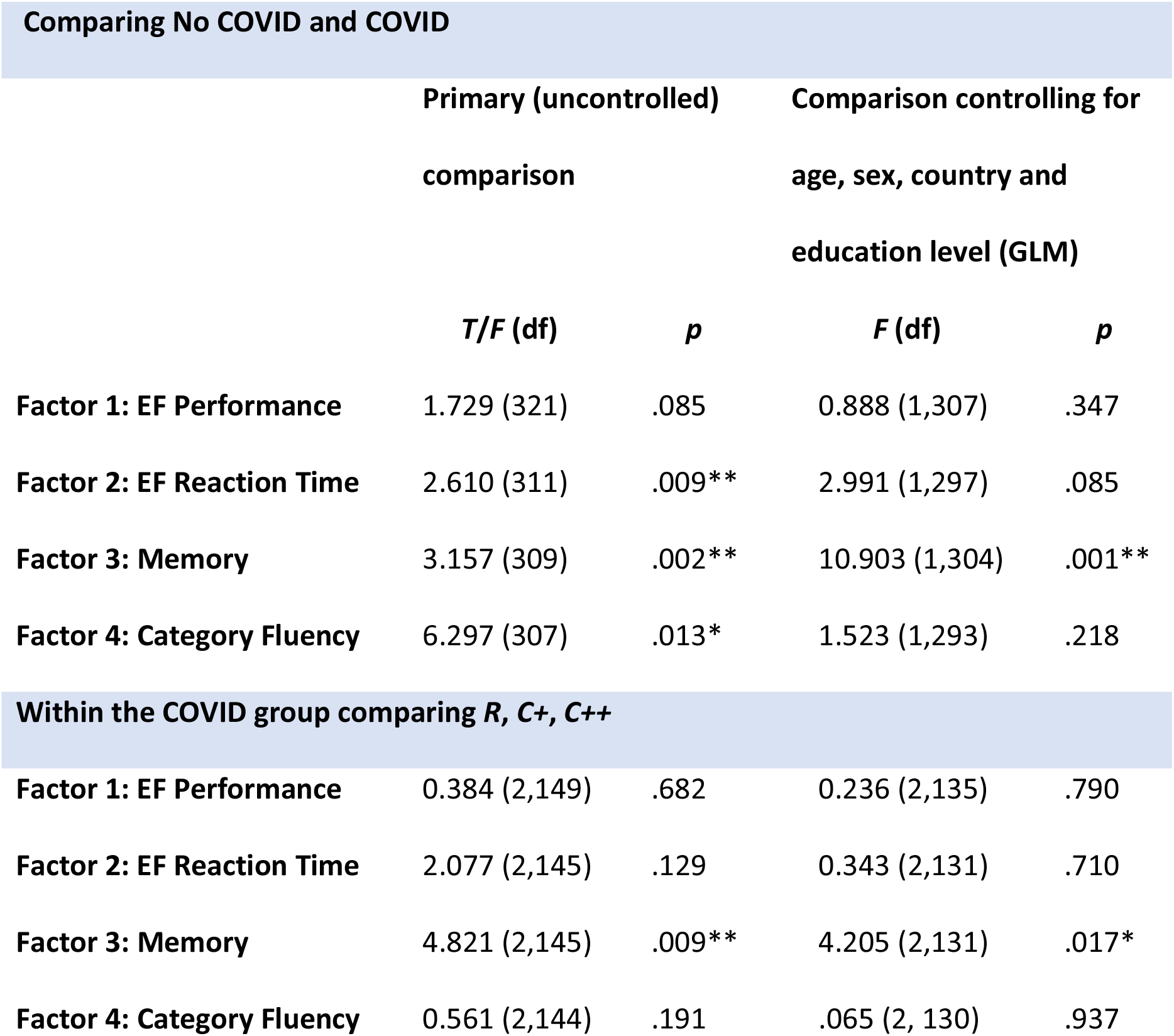
Cognitive performance factors across COVID and No COVID groups (top) and symptom severity levels (bottom).

| Comparing No COVID and COVID |  |  |  |  |
| --- | --- | --- | --- | --- |
|  | Primary (uncontrolled)<br>comparison |  | Comparison controlling for<br>age, sex, country and<br>education level (GLM) |  |
|  | *T/F (df)* | *p* | *F (df)* | *p* |
| Factor 1: EF Performance | 1.729 (321) | .085 | 0.888 (1,307) | .347 |
| Factor 2: EF Reaction Time | 2.610 (311) | .009** | 2.991 (1,297) | .085 |
| Factor 3: Memory | 3.157 (309) | .002** | 10.903 (1,304) | .001** |
| Factor 4: Category Fluency | 6.297 (307) | .013* | 1.523 (1,293) | .218 |
| Within the COVID group comparing *R, C+, C++* |  |  |  |  |
| Factor 1: EF Performance | 0.384 (2,149) | .682 | 0.236 (2,135) | .790 |
| Factor 2: EF Reaction Time | 2.077 (2,145) | .129 | 0.343 (2,131) | .710 |
| Factor 3: Memory | 4.821 (2,145) | .009** | 4.205 (2,131) | .017* |
| Factor 4: Category Fluency | 0.561 (2,144) | .191 | .065 (2, 130) | .937 |

**Figure 2.**
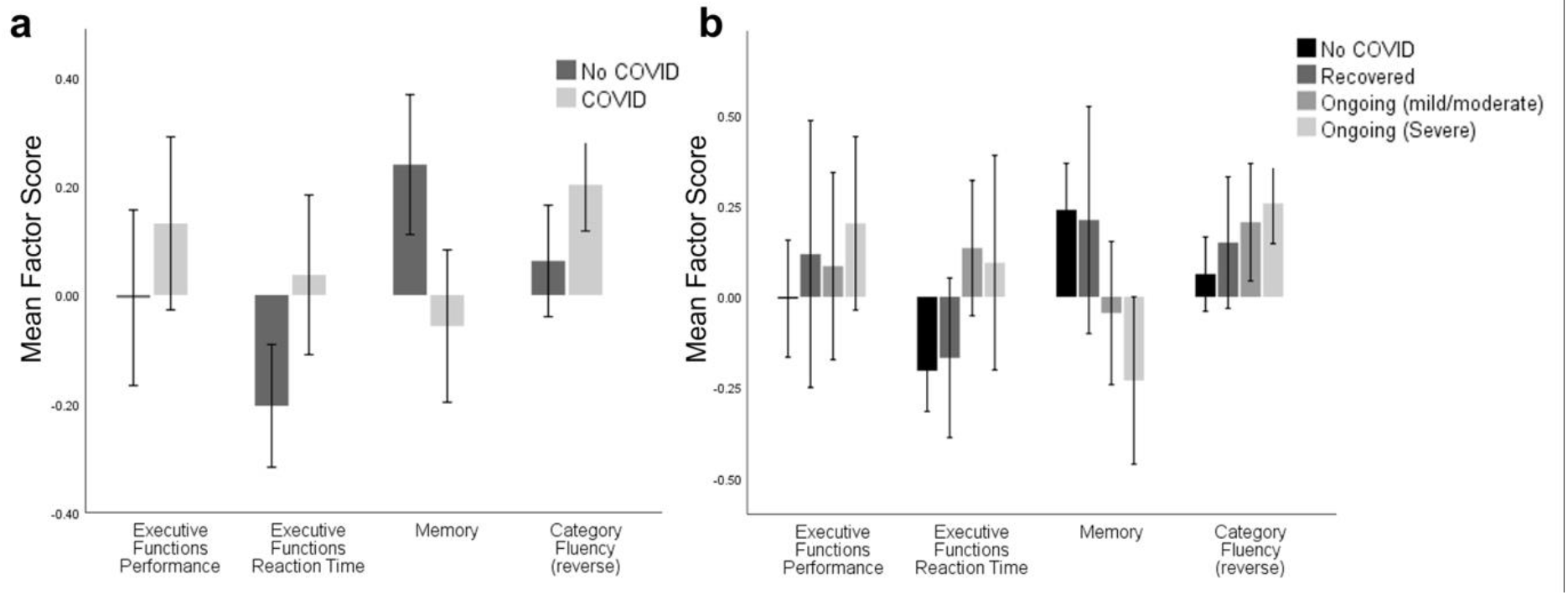
Cognitive factor scores across the No COVID (a) COVID group and (b) the three ongoing severity groups. Significant differences were seen between the No COVID group and Ongoing (Mild/Moderate) on Memory (*t*(87.6) = 2.4, *p* = .018), and between No COVID and Ongoing (Severe) on Memory (*t*(99.8) = 3.9, *p* < .001) and Category Fluency (*t*(152) = 3.05, *p* < .003). After controlling for demographic variables, only the differences in Memory maintained significance (see supplementary Table 2). Error bars: +/− 2 SE.

For individual variables, primary analysis suggested that individuals who had experienced COVID-19 infection had significantly lower performance (*U* = 3.29, *p* < .001) and slower reaction time (*U* = 3.53, *p* < .001) than the No COVID group on the Word List Recognition Memory Test (Table 2). After controlling for age, sex, country and education level, these effects were maintained (% correct: *F*(1,315) = 6.77, *p* = .01; RT: F(1,315) = 12.66, *p* < .001)), with d’ becoming significant (*F*(1,315) = 5.78, *p* = .017). A much weaker trend was seen in the Pictorial Associative Memory Test, suggesting a reduced performance in the COVID group (*t* = 1.91, *p* = .056) and no impact on reaction time (*p* = .671). When controlling for age, sex, country and education level, the significance of this group effect strengthened, suggesting that those who had experienced COVID-19 infection scored lower than the No COVID group (*F*(1,319) = 4.01, *p* = .046). Considering only analyses controlling for demographic factors, only reaction time on the Word List Recognition survived conservative correction for multiple comparisons (Sidak *α* = .0028).

**Table 2.**
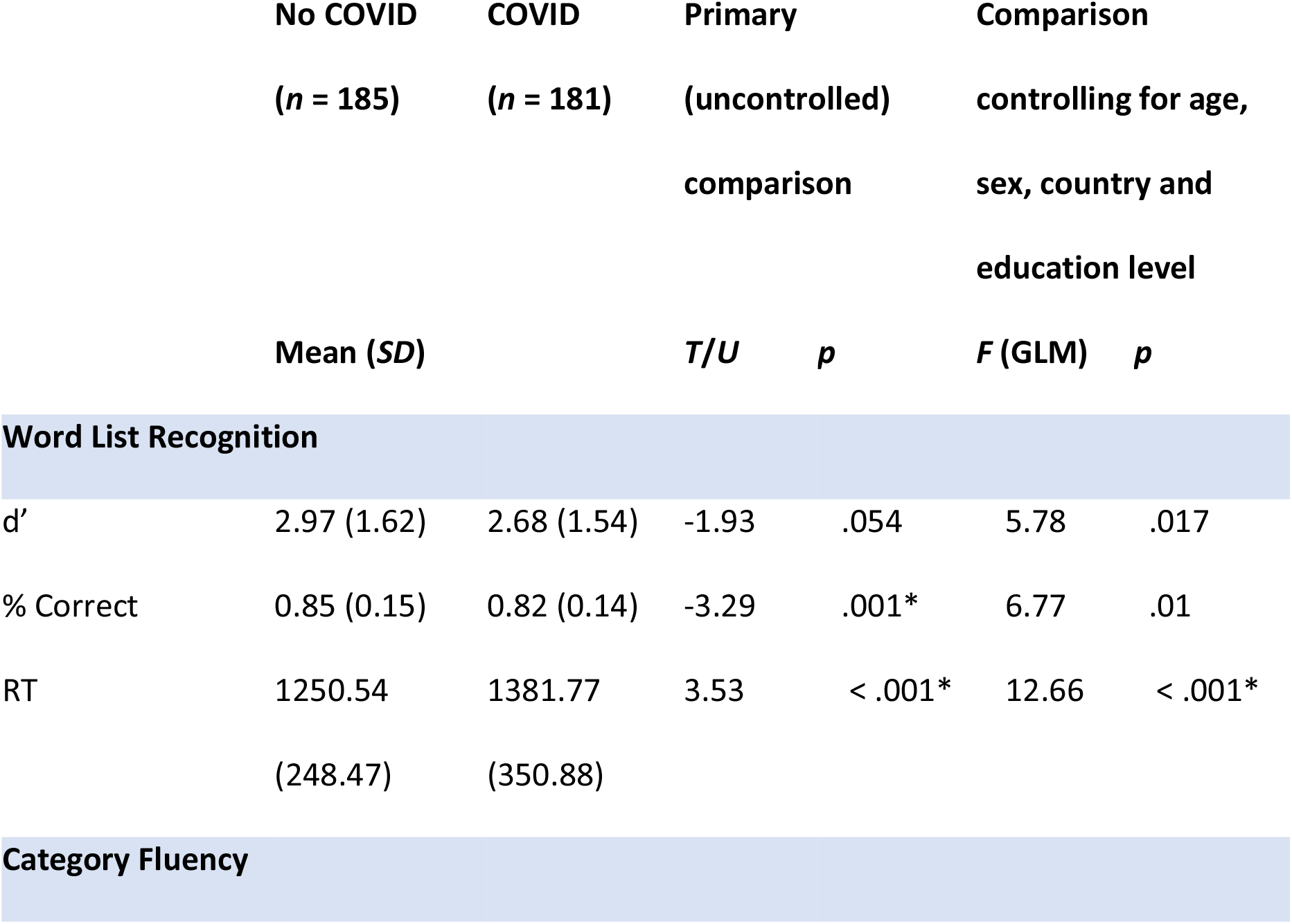
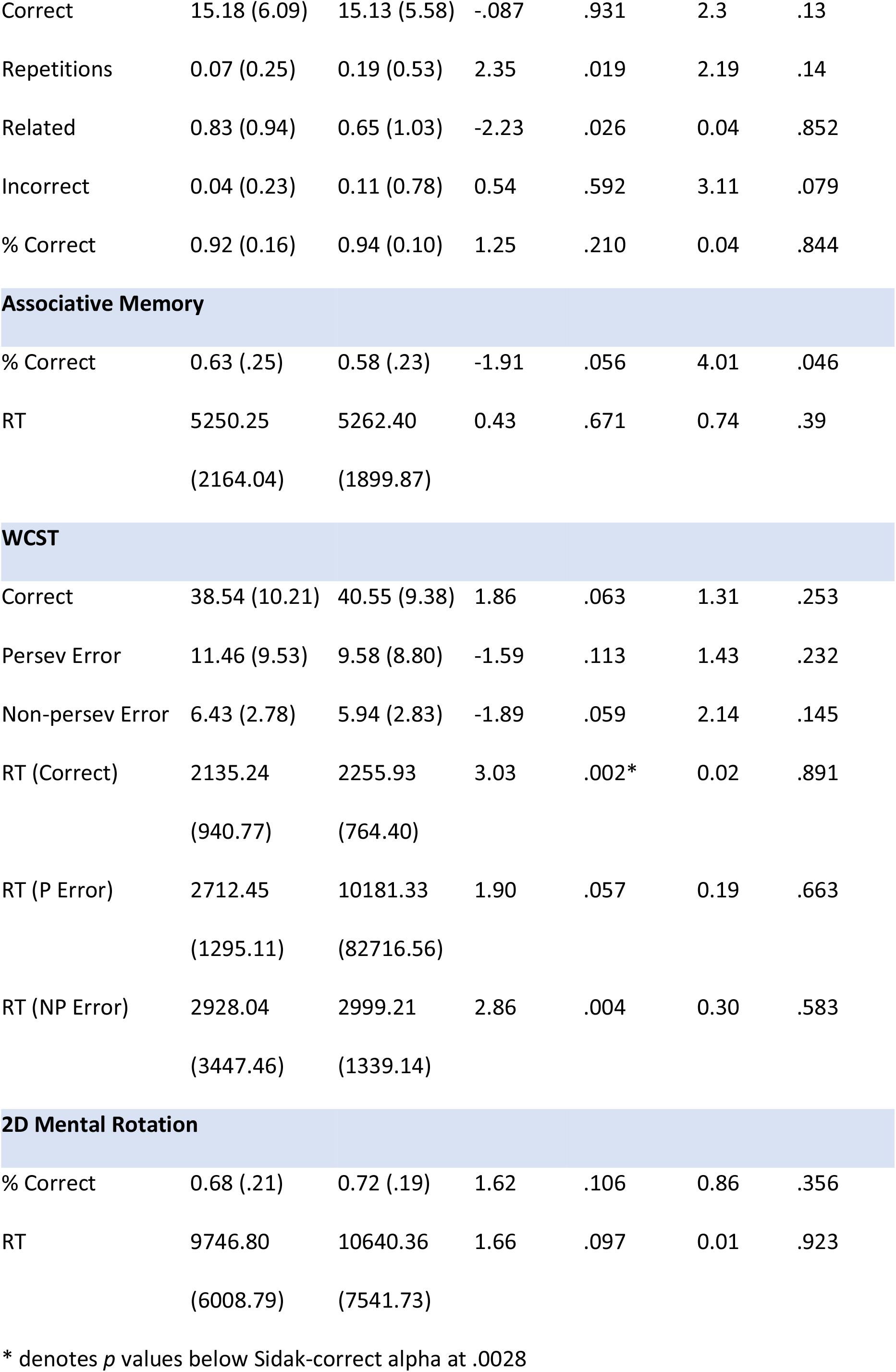
Cognitive task results between No COVID and COVID groups.

|  | No COVID<br>(*n* = 185) | COVID<br>(*n* = 181) | Primary<br>(uncontrolled)<br>comparison |  | Comparison<br>controlling for age,<br>sex, country and<br>education level |  |
| --- | --- | --- | --- | --- | --- | --- |
|  | Mean (*SD*) |  | *T/U* | *p* | *F* (GLM) | *p* |
| **Word List Recognition** |  |  |  |  |  |  |
| *d'* | 2.97 (1.62) | 2.68 (1.54) | -1.93 | .054 | 5.78 | .017 |
| % Correct | 0.85 (0.15) | 0.82 (0.14) | -3.29 | .001* | 6.77 | .01 |
| RT | 1250.54<br>(248.47) | 1381.77<br>(350.88) | 3.53 | < .001* | 12.66 | < .001* |
| **Category Fluency** |  |  |  |  |  |  |
| Correct | 15.18 (6.09) | 15.13 (5.58) | -.087 | .931 | 2.3 | .13 |
| Repetitions | 0.07 (0.25) | 0.19 (0.53) | 2.35 | .019 | 2.19 | .14 |
| Related | 0.83 (0.94) | 0.65 (1.03) | -2.23 | .026 | 0.04 | .852 |
| Incorrect | 0.04 (0.23) | 0.11 (0.78) | 0.54 | .592 | 3.11 | .079 |
| % Correct | 0.92 (0.16) | 0.94 (0.10) | 1.25 | .210 | 0.04 | .844 |
| Associative Memory |  |  |  |  |  |  |
| % Correct | 0.63 (.25) | 0.58 (.23) | -1.91 | .056 | 4.01 | .046 |
| RT | 5250.25<br>(2164.04) | 5262.40<br>(1899.87) | 0.43 | .671 | 0.74 | .39 |
| WCST |  |  |  |  |  |  |
| Correct | 38.54 (10.21) | 40.55 (9.38) | 1.86 | .063 | 1.31 | .253 |
| Persev Error | 11.46 (9.53) | 9.58 (8.80) | -1.59 | .113 | 1.43 | .232 |
| Non-persev Error | 6.43 (2.78) | 5.94 (2.83) | -1.89 | .059 | 2.14 | .145 |
| RT (Correct) | 2135.24<br>(940.77) | 2255.93<br>(764.40) | 3.03 | .002* | 0.02 | .891 |
| RT (P Error) | 2712.45<br>(1295.11) | 10181.33<br>(82716.56) | 1.90 | .057 | 0.19 | .663 |
| RT (NP Error) | 2928.04<br>(3447.46) | 2999.21<br>(1339.14) | 2.86 | .004 | 0.30 | .583 |
| 2D Mental Rotation |  |  |  |  |  |  |
| % Correct | 0.68 (.21) | 0.72 (.19) | 1.62 | .106 | 0.86 | .356 |
| RT | 9746.80<br>(6008.79) | 10640.36<br>(7541.73) | 1.66 | .097 | 0.01 | .923 |

\* denotes *p* values below Sidak-correct alpha at .0028

For Category Fluency, uncorrected analysis found that although the COVID group repeated more words (*U* = 2.35, *p* = .019), they gave fewer incorrect (related) words (*U* = 2.23, *p* = .026) than the No COVID group. However, these effects disappeared after factoring out age, sex, country and education level.

#### 3.2.2 Other Tasks

There were no significant differences between the groups on the Executive Function Performance factor, but there was a significant group different in Executive Function Reaction Time (*t*(311) = 2.610, *p* = .009), but this dropped below significance once age, sex, country and education were accounted for (see Table 1, Figure 2).

In terms of individual variables, there were no group differences in performance on the WCST, however the COVID-group had significantly slower reaction time on trials with both correct responses (*U* = 3.03, *p* = .002; see Table 2) and non-perseverative errors (*U* = 2.86, *p* = .004). No significant difference was found after controlling for age, sex, country and education level. There were no significant differences between groups on performance on the 2D Mental Rotation Test.

### 3.3 Ongoing Symptom-Severity and Cognition

#### 3.3.1 Memory and Word Finding

There was a significant difference between ongoing symptom severity groups in the memory factor (*F*(2,150) = 5.724, *p* = .004) which was weakened but still significant when demographic factors were accounted for (*F*(2,136) = 3.653, *p* = .028). Pairwise analysis controlling for demographic variables showed a significant difference between the Recovered and Ongoing (severe) groups (*F*(1,88) = 6.414, *p* = .013). There was no association between symptom severity and the Category Fluency factor (see Table 1).

In terms of individual variables (see Table 3), significant differences between ongoing symptom sub-groups were found on Word List % correct (*H*(3) = 22.51, *p* < .001; Figure 3) and reaction time (*H*(3) = 24.07, *p* < .001). Pairwise tests with Sidak *α* = .008 revealed that those with severe ongoing symptoms had lower % correct than the No COVID group (*p* < .001) and those that had recovered (*p* < .001), and had slower reaction time than the No COVID group (*p* < .001). Those with mild-moderate ongoing symptoms also had slower reaction time than the No COVID group (*p* < .001) and Recovered group (*p* = .004). When age, sex, country and education level were factored out by general linear model (GLM), d’ (*F*(3,310) = 2.90, *p* = .035), % correct (*F*(3,310) = 4.99, *p* = .002), and reaction time (*F*(3,310) = 6.88, *p* < .001) differences were all significant, but only % correct and reaction time survived correction for multiple comparisons (Sidak *α* = .0028). Pairwise tests suggested that those with severe ongoing symptoms had significant lower d’ (*p* = .004), lower % correct (*p* < .001), and slower reaction time than the No COVID group (*p* < .001). Those with mild-moderate ongoing symptoms still had slower reaction time than the No COVID group (*p* < .001). In contrast to these findings with Word List Recognition Memory, primary analysis did not find significant group differences on Pictorial Associative Memory—on either performance or reaction time. However, after controlling age, sex, country and education level, a main effect emerged for % correct (*F*(3,314) = 2.94, *p* = .034); however this did not survive correction for multiple comparisons (Sidak *α* = .0028). Nonetheless, pairwise comparisons suggested that those with severe ongoing symptoms scored lower than the No COVID group (*p* = .005, Sidak *α* = .008).

**Table 3.**
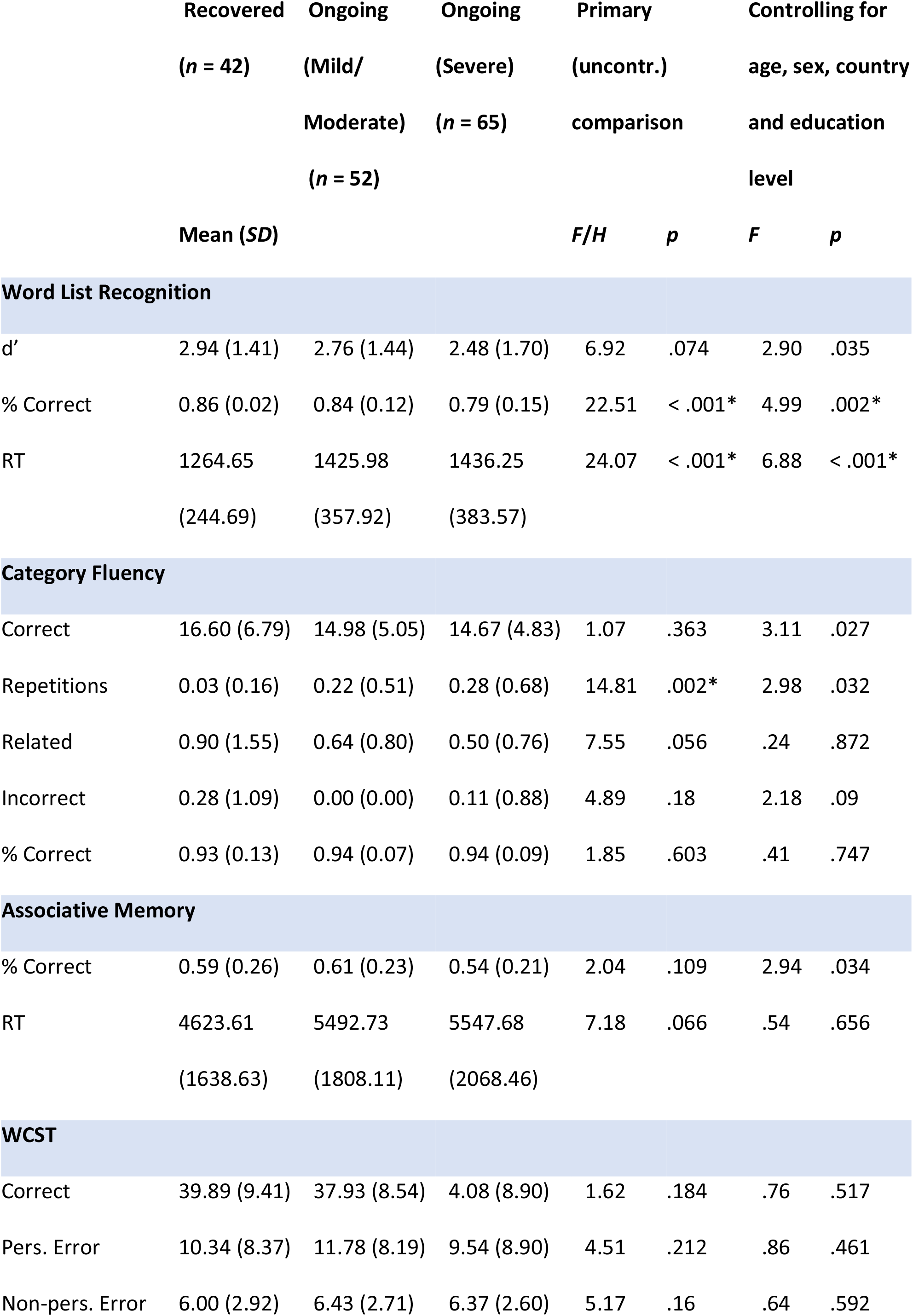
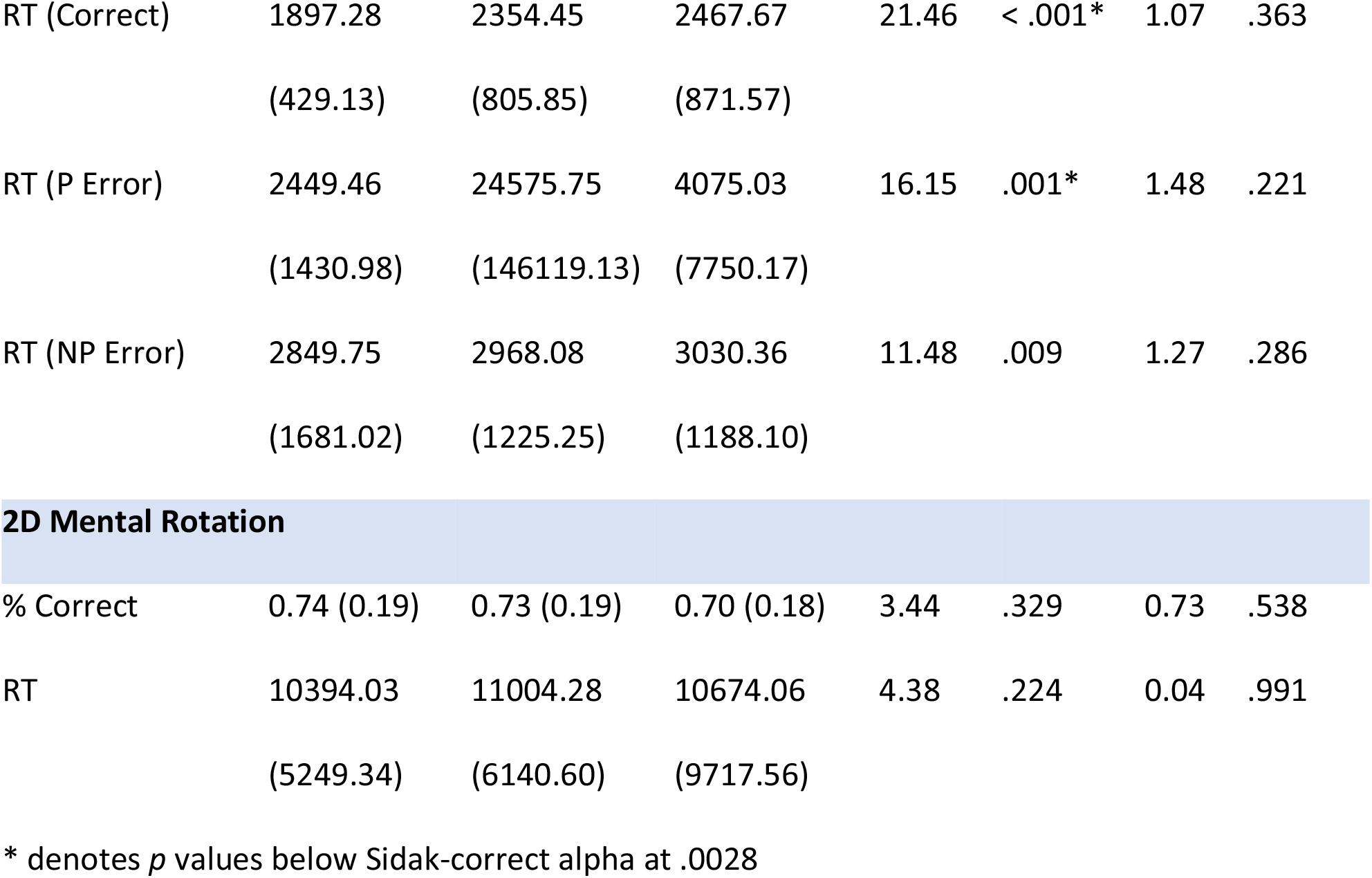
Cognitive task results among Recovered, Ongoing (Mild/Moderate) and Ongoing (Severe) groups.

|  | Recovered<br>(*n* = 42) | Ongoing<br>(Mild/<br>Moderate)<br>(*n* = 52) | Ongoing<br>(Severe)<br>(*n* = 65) | Primary<br>(uncontr.)<br>comparison | Controlling for<br>age, sex, country<br>and education<br>level |  |  |
| --- | --- | --- | --- | --- | --- | --- | --- |
|  | Mean (*SD*) |  |  | *F/H* | *p* | *F* | *p* |
| **Word List Recognition** |  |  |  |  |  |  |  |
| *d'* | 2.94 (1.41) | 2.76 (1.44) | 2.48 (1.70) | 6.92 | .074 | 2.90 | .035 |
| % Correct | 0.86 (0.02) | 0.84 (0.12) | 0.79 (0.15) | 22.51 | < .001* | 4.99 | .002* |
| RT | 1264.65<br>(244.69) | 1425.98<br>(357.92) | 1436.25<br>(383.57) | 24.07 | < .001* | 6.88 | < .001* |
| **Category Fluency** |  |  |  |  |  |  |  |
| Correct | 16.60 (6.79) | 14.98 (5.05) | 14.67 (4.83) | 1.07 | .363 | 3.11 | .027 |
| Repetitions | 0.03 (0.16) | 0.22 (0.51) | 0.28 (0.68) | 14.81 | .002* | 2.98 | .032 |
| Related | 0.90 (1.55) | 0.64 (0.80) | 0.50 (0.76) | 7.55 | .056 | .24 | .872 |
| Incorrect | 0.28 (1.09) | 0.00 (0.00) | 0.11 (0.88) | 4.89 | .18 | 2.18 | .09 |
| % Correct | 0.93 (0.13) | 0.94 (0.07) | 0.94 (0.09) | 1.85 | .603 | .41 | .747 |
| **Associative Memory** |  |  |  |  |  |  |  |
| % Correct | 0.59 (0.26) | 0.61 (0.23) | 0.54 (0.21) | 2.04 | .109 | 2.94 | .034 |
| RT | 4623.61<br>(1638.63) | 5492.73<br>(1808.11) | 5547.68<br>(2068.46) | 7.18 | .066 | .54 | .656 |
| **WCST** |  |  |  |  |  |  |  |
| Correct | 39.89 (9.41) | 37.93 (8.54) | 4.08 (8.90) | 1.62 | .184 | .76 | .517 |
| Pers. Error | 10.34 (8.37) | 11.78 (8.19) | 9.54 (8.90) | 4.51 | .212 | .86 | .461 |
| Non-pers. Error | 6.00 (2.92) | 6.43 (2.71) | 6.37 (2.60) | 5.17 | .16 | .64 | .592 |
| RT (Correct) | 1897.28<br>(429.13) | 2354.45<br>(805.85) | 2467.67<br>(871.57) | 21.46 | < .001* | 1.07 | .363 |
| RT (P Error) | 2449.46<br>(1430.98) | 24575.75<br>(146119.13) | 4075.03<br>(7750.17) | 16.15 | .001* | 1.48 | .221 |
| RT (NP Error) | 2849.75<br>(1681.02) | 2968.08<br>(1225.25) | 3030.36<br>(1188.10) | 11.48 | .009 | 1.27 | .286 |
| 2D Mental Rotation |  |  |  |  |  |  |  |
| % Correct | 0.74 (0.19) | 0.73 (0.19) | 0.70 (0.18) | 3.44 | .329 | 0.73 | .538 |
| RT | 10394.03<br>(5249.34) | 11004.28<br>(6140.60) | 10674.06<br>(9717.56) | 4.38 | .224 | 0.04 | .991 |

\* denotes $p$ values below Sidak-correct alpha at .0028

**Figure 3.**
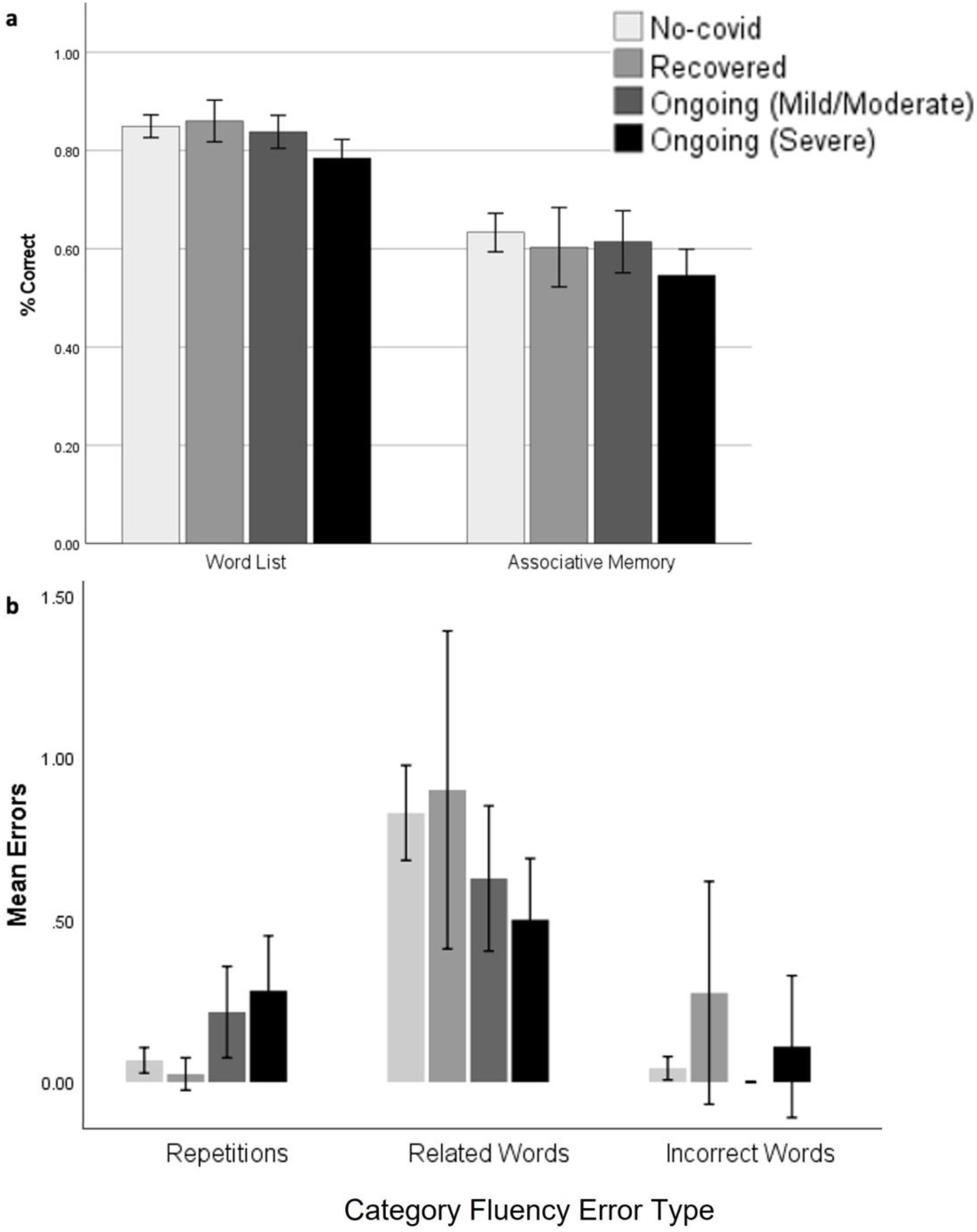
Word List and Associative Memory performance across ongoing symptom groups (top); Category Fluency errors across groups on ongoing symptom severity (bottom). Error bars: +/− 2 SE.

For Category Fluency, primary analysis showed a significant group effect in repetitions (*H*(3) = 14.81, *p* = .002; Figure 3). Pairwise comparison with Sidak *α* = .008 found that those with severe ongoing symptoms had more repeated words than both the No COVID (*p* = .002) and Recovered groups (*p* = .004). When GLM controlling for age, sex, country and education level was conducted, there were significant main effects on number of correct words (*F*(3,301) = 3.11, *p* = .027) and repetitions (*F*(3,301) = 2.98, *p* = .032). but neither of these survived correction for multiple comparisons (Sidak *α* = .0028). Pairwise tests showed that those with severe ongoing symptoms had fewer correct words than the Recovered group (*p* = .008), but no pairwise comparisons were significant for repetitions.

#### 3.3.2 Other Tasks

There was no effect of symptom severity group on either of the Executive Function factors (see Table 1). There were significant group effects for the WCST in reaction time for trials with correct responses (*H*(3) = 21.46, *p* < .001), perseverative (*H*(3) = 16.15, *p* = .001), and non-perseverative errors (*H*(3) = 11.48, *p* = .009). Pairwise tests with Sidak *α* = .008 showed that those with mild/moderate ongoing symptoms had slower reaction time for trials with correct responses than the No COVID group (*p* = .005) and Recovered group (*p* = .008). Similarly, those with severe ongoing symptoms were slower for correct responses than those who recovered (*p* < .001) and the No COVID group (*p* < .001). For trials containing perseverative errors, both those with mild-moderate (*p* = .002) and severe (*p* = .002) ongoing symptoms has slower reaction times than those who recovered. Those with mild-moderate ongoing symptoms were also slower than the No COVID group for trials containing non-perseverative errors (*p* = .005). However, after controlling for age, sex, country and education level, all these significances disappeared. There were no significant effects in 2D Mental Rotation.

### 3.4 Initial Illness and Subsequent Cognitive Performance

#### 3.4.1 Severity of Initial Illness

We assessed whether more severe initial illness (grouped into three: Asymptomatic/very mild; Mild (bed-bound); Moderate/severe (very ill or hospitalized)) was associated with cognitive performance at the time of test (often weeks or months later). First we examined this in terms of the cognitive task factors. There was no effect of initial symptom severity on any of the cognitive task factor scores (EF Performance: *F*(2,149) = .479, *p* = .620; EF RT: *F*(2,146) = .019, *p* = .982; Memory: *F*(2,146) = 1.087, *p* = .340; Category Fluency: *F*(1,145) = 1.171, *p* = .313).

Next, we examined which (if any) individual cognitive task variables could be associated with variance in initial illness severity (asymptomatic/very mild, mild, moderate/severe) using independent simple regression models with COVID-19 illness severity as the dependent variable and all cognitive task variable as predictors. There was a significant association for Word List Recognition (*F*(1,142) = 6.369, *p* = .013, standardized *B* = −0.207, Adjusted *R^2^* = .036), but not other cognitive task was associated with initial illness severity. These associations did not survive correction for multiple comparisons (Sidak *α* = .0028).

We also examined whether any particular diagnoses during the initial illness was related to subsequent cognitive performance. After removing diagnoses with very low prevalence (< than 4%), none of the remaining diagnoses (hypoxia, blood clots, and Inflammatory syndrome) presented any significant association with cognitive performance.

#### 3.4.2 Nature of Initial illness and Cognitive Performance

##### 3.4.2.1 Individual Neurological Symptoms

To test whether any of specific neurological symptoms experienced during the first 3 weeks of illness (initial symptoms) were related to subsequent cognitive performance, we carried out multiple linear regressions with cognitive performance factors as the dependent variable, and the neurological symptoms as possible predictors. Almost no participants showed hallucination or delirium (< 10% of participants), so these were removed from analysis.

A single early neurological symptom emerged as predicting variance in cognitive factors. Both Executive Function Performance (*ή_p_^2^* = .03) and Memory (*ή_p_^2^* = .038) were predicted by initial disorientation (EF Performance: *R_adj_^2^* = .024, *p* = .032; Memory: *R_adj_^2^* = .031, *p* = .017). Variance in Executive Function RT and Category Fluency factors were not predicted by early neurological symptoms. With individual cognitive tests as the dependent variable, several models emerged, however, none of survived correction for multiple comparisons (Sidak *α* = .0028; Supplementary Table 3). Headache severity was associated with slowed reaction time of the Word List Recognition Test (*p* = .005), and fewer correct answers on the Category Fluency (*p* = .003) and Pictorial Associative Memory (*p* = .036) Tests. Confusion predicted the percentage of correct answers of the Category Fluency (*p* = .047) and the Word List Recognition Tests (*p* = .006). Altered consciousness predicted Word List Recognition d’ (*p* = .003), and dizziness predicted perseverative errors in the WCST (*p* = .035). Disorientation predicted WCST correct answers (*p* = .019), and numbness predicted WCST reaction time for trials with correct answers (*p* = .003). Speech difficulty, disturbed vision, and loss of smell/taste did not predict any cognitive outcome.

##### 3.4.2.2 Initial Symptom Factors

As reported in our previous publication with this sample (Guo et al. 2021), we used exploratory factor analysis to reduce reported symptoms into related factors. For initial symptoms, 5 factors were identified: “Neurological” (characterized by disorientation, delirium and visual disturbances); “Fatigue/Systemic” (characterized by fatigue, chest pain/tightness and muscle/body pains); “Gastrointestinal” (characterized by diarrhea, vomiting and nausea); “Respiratory/Infectious” (characterized by fever, cough, and breathing issues) and “Dermatological” (characterized by itchy welts, rash and foot sores). To assess whether any of the symptom-factors predicted any aspect of the different cognitive tasks, we conducted various multiple linear regression models (backward elimination method) with the symptom factors as predictors and cognitive task as the dependent variables.

No model significantly predicted variation in the EF Performance or Category Fluency factors. Individual differences in EF Reaction Time were significantly predicted by a model which contained only the Dermatological factor (*ή_p_^2^* = .079) and predicted over 8% of variance (*R_adj_^2^* = .081, *p* < .001; see Figure 4). Individual differences on the Memory factor were significantly predicted by a model containing the Fatigue/Systemic factor (*ή_p_^2^* = .061) and predicted 5.4% of variance (*R_adj_^2^* = .054, *p* = .002).

**Figure 4.**
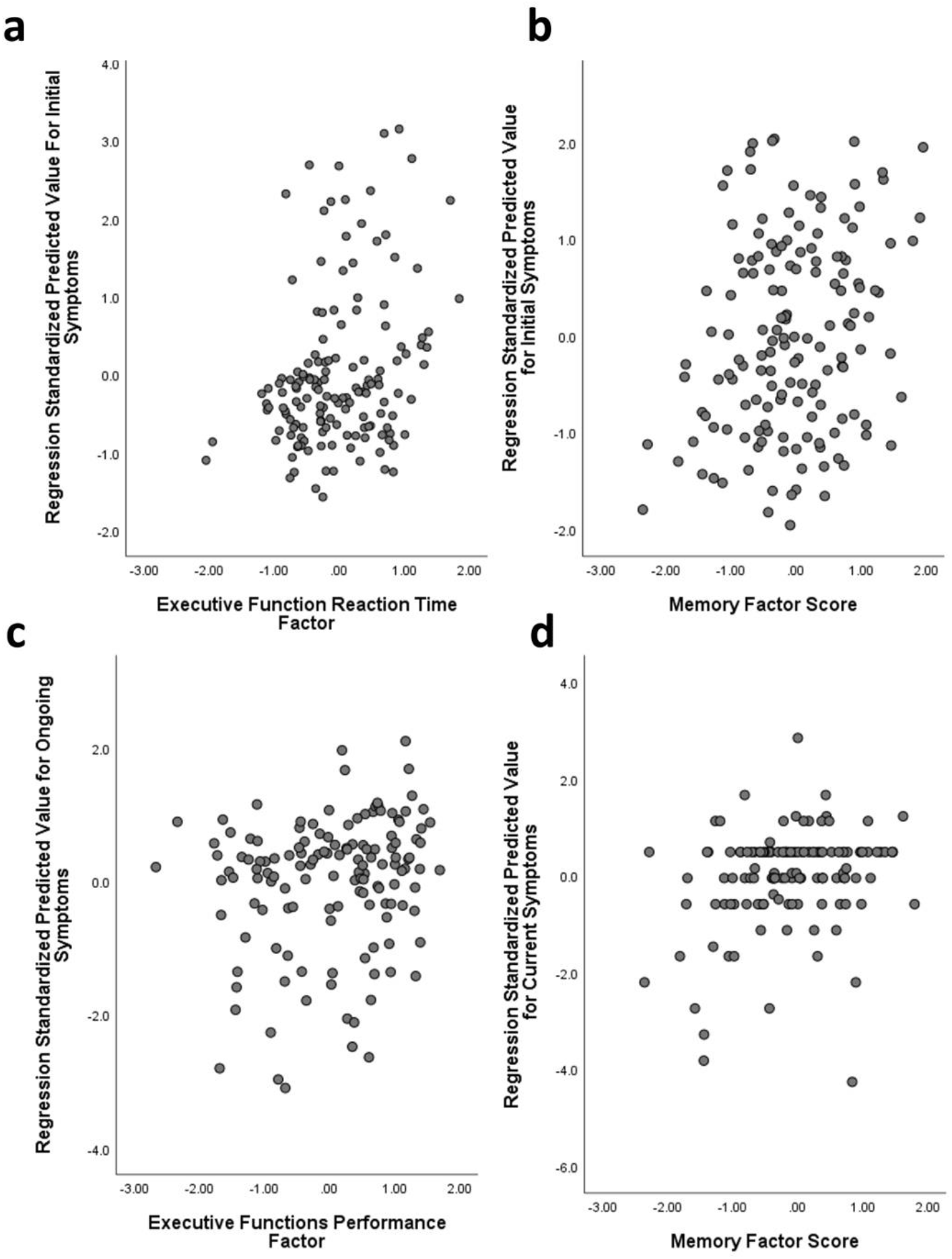
Symptom factors predicting cognitive task factors. A) Initial symptoms model (Dermatological) predicting EF Reaction Time; b) Initial symptoms model (Fatigue/Systemic) predicting Memory; c) Ongoing symptoms model (Neurological) predicting EF Performance and d) Current symptoms model (Neurological) predicting Memory. Note that symptom factors are reverse coded (lower numbers translate to more severe symptoms).

The initial-symptom factors predicted aspects of all the individual cognitive tasks (Table 4). The Fatigue/Systemic factor predicted d’ (*p* = .008) and reaction time (*p* = .003) within the Word List Recognition Test, as well as Category Fluency correct answers (*p* = .014). The Fatigue/Systemic factor also predicted WCST reaction time (for correct answers, *p* = .002) in combination with the Dermatological factor. When the Fatigue/Systemic factor was combined with Respiratory/Infectious factor, significant variance was predicted in Word List % correct (*p* = .003), and the Respiratory/Infectious factor independently predicted correct choices on the WCST (*p* = .042). Finally, the Dermatological factor independently predicted reaction time in the 2D Mental Rotation Test (*p* = .001) and the Pictorial Associative Memory Test (*p* = .048). Compared against a corrected alpha (Sidak *α* = .0028), the models predicting WCST reaction time and 2D Mental Rotation maintained significance (see Table 4).

**Table 4.**
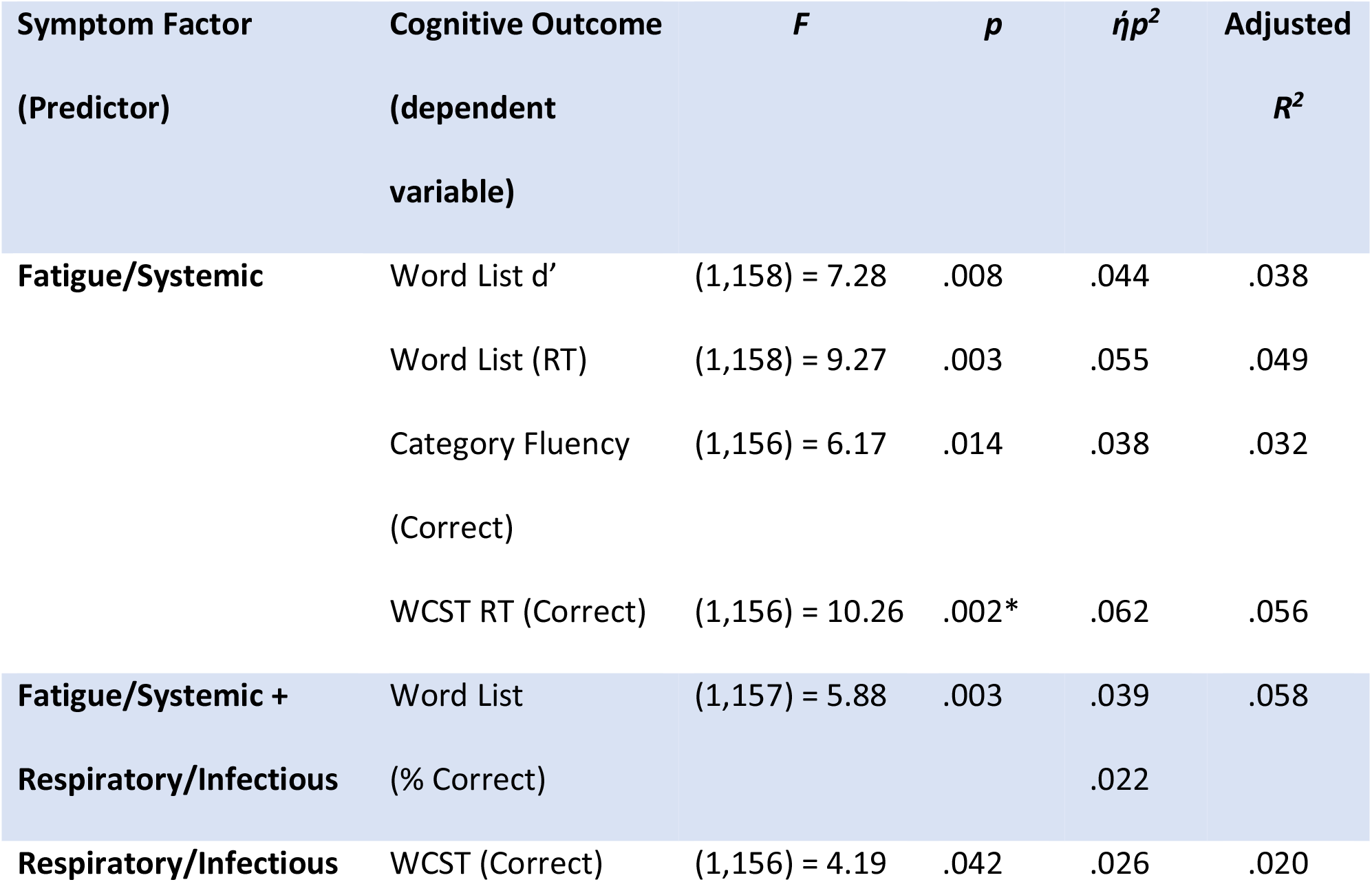
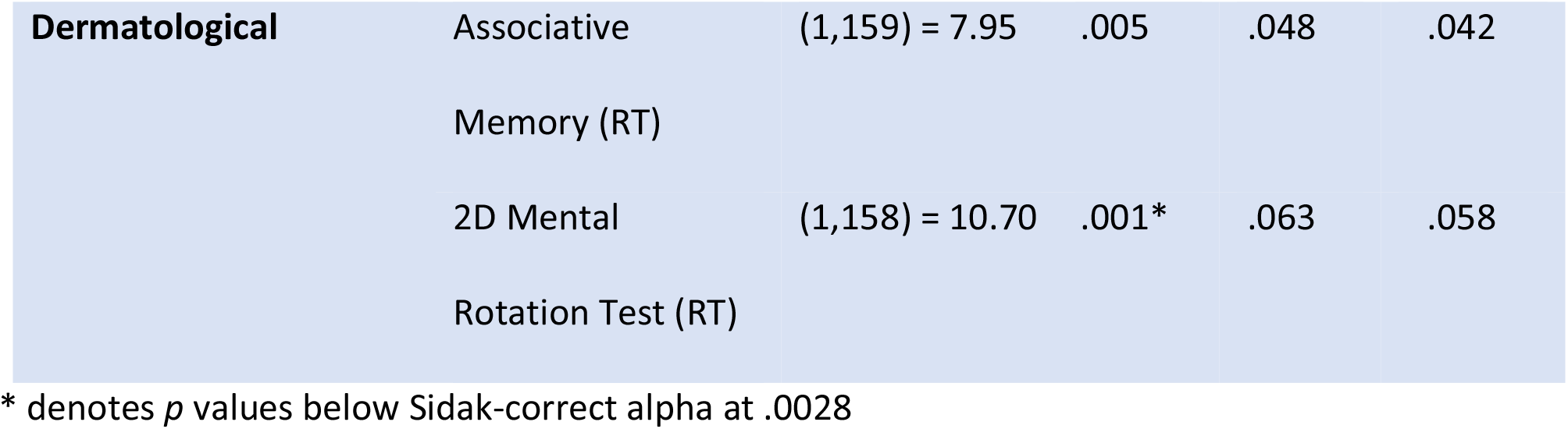
Initial symptom factors and subsequent cognitive performance.

### 3.5 Nature of Ongoing Illness and Cognitive Performance

#### 3.5.1 Ongoing Symptoms and Cognitive Performance

As reported in our previous publication with this sample (Guo et al. 2021), 6 factors were identified within the ongoing symptoms: “Neurological” (characterized by disorientation, confusion and delirium); “Gastrointestinal/Autoimmune” (characterized by diarrhea, hot flushes, and nausea); “Cardio-Pulmonary” (characterized by breathing issues, chest pain/tightness and fatigue); “Dermatological/Fever” (characterized by face/lips swelling, foot sores and itchy welts); “Mood” (characterized by depression, anxiety and vivid dreams) and “Appetite Loss” (characterized by weight loss and loss of appetite). To assess whether symptoms experienced in the time since the initial infection predicted any aspect of the different cognitive tasks, we entered the ongoing symptom factors into a series of regressions with the cognitive task variables as dependents.

For these ongoing symptoms, no model significantly predicted variance in the EF Reaction Time, Memory or Category Fluency factors. The Neurological factor alone predicted variance in EF Performance (*ή_p_^2^* = .031; *R_adj_^2^* = .024, *p* = .037). Different symptom factors were able to explain variance in different individual cognitive tasks (Table 5). The Cardio-Pulmonary factor predicted a significant amount of variance in Word List % correct (*p* = .03) and reaction time in WCST trials containing correct answers (*p* = .01). The Neurological factor predicted variance in WCST correct answers (*p* = .46), and in combination with the Dermatological/Fever factor predicted performance on the WCST (*p* = .013). The Neurological factor and mood factors together predicted % of words produced that were correct in the Category Fluency Test (*p* = .004) Finally, the Gastrointestinal/Autoimmune factor predicted variation in Word List reaction time (*p* = .046). None of these associations survived correction for multiple comparisons (Sidak *α* = .0028).

**Table 5.**
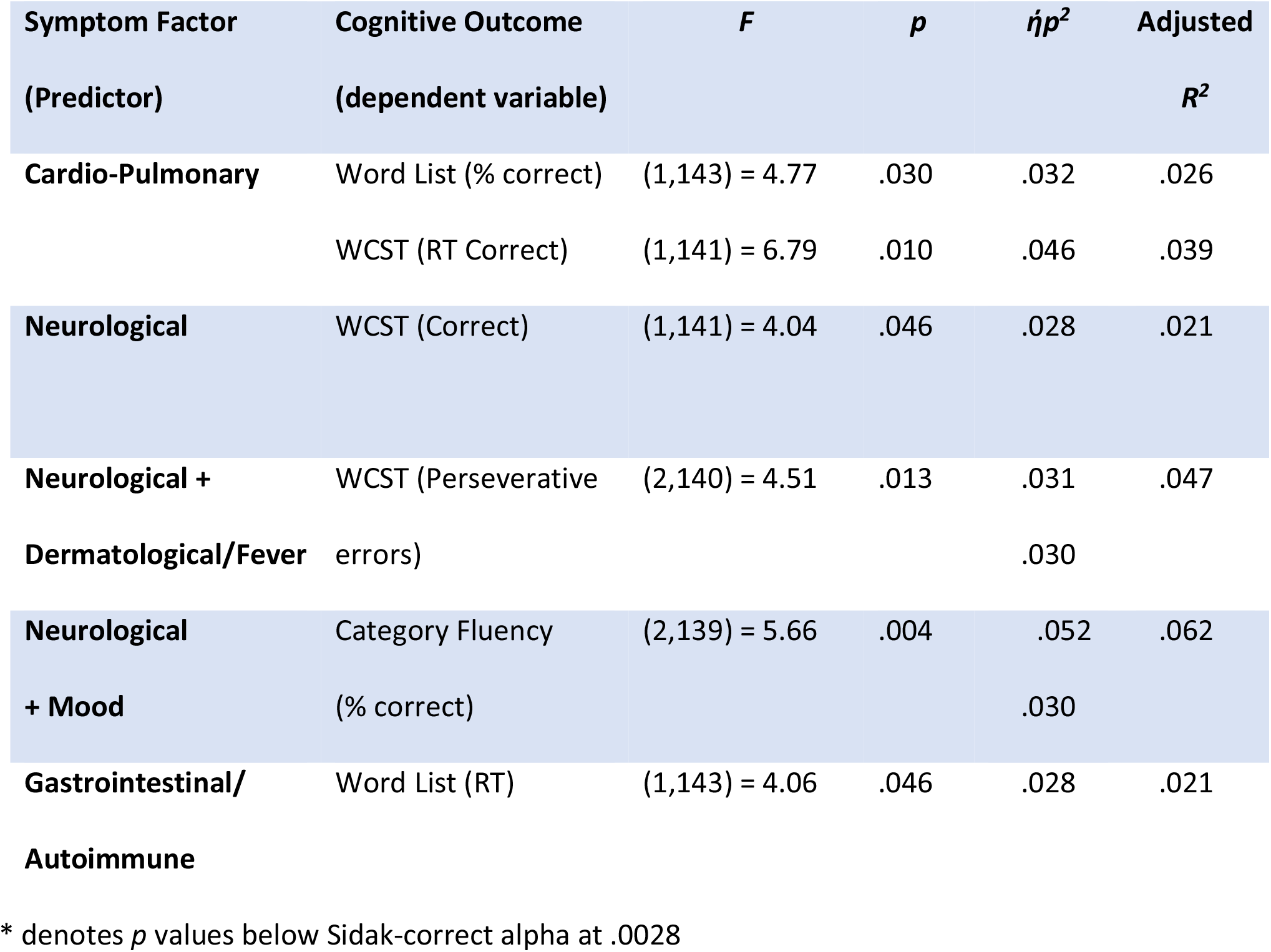
Ongoing symptom factors and subsequent cognitive performance.

### 3.6 Nature of Current Illness and Cognitive Performance

#### 3.6.1 Severity of Current Illness

Given the often-cyclical nature of symptoms, participants were asked to report to what degree they were experiencing a “bad day” in terms of symptoms on the day of testing. To address the question of whether group differences in performance were due to severity of illness on the day of testing, we first assessed whether completing the test on a “bad day” impacted cognitive performance. No cognitive factor showed any significant associations with current symptom severity. In terms of individual cognitive task variables, there were group effects in Category Fluency repetitions (*F*(4,117) = 5.809, *p* < .001), 2D Mental Rotation reaction time (*F*(4,118) = 5.371, *p* = .001), both of which survived correction for multiple comparisons (Sidak *α* = .0028). However, no effect was directional (with the only significant correlation being with 2D Mental Rotation performance (*r* = −.184, *p* = .042, which did not survive correction for multiple comparisons (Sidak *α* = .0028)).

To test whether associations between ongoing symptoms and cognitive performance were not better explained by the symptoms’ severity on the day of testing, rather than the presence of ongoing symptoms per se, we performed stepwise regressions with the cognitive factors as the dependent, current symptom severity (good/bad day) as the first step and ongoing symptom subgroup (*R*/*C+*/*C++*) as the second step. Current symptom severity was not a significant predictor of any cognitive outcome.

#### 3.6.2 Current Symptom Factors

As reported in our previous publication (Guo et al. 2021), factor scores for current symptoms were calculated from the 6 ongoing symptom factors. No current symptom factors significantly predicted individual differences in either Executive Function factors or the Category Fluency factor. A model containing the neurological factor (*ή_p_^2^* = .041) predicted variance in the Memory factor (*R_adj_^2^* = .034, *p* = .018).

In terms of individual task variables, the degree to which current symptoms aligned with the Mood factor (*ή_p_^2^* = .043) predicted the percentage of correct words in the Category Fluency Test (*R_adj_^2^* = .037, *p* = .013), while alignment with the Dermatological/Fever factor (*ή_p_^2^* = .029) predicted variance in the number of repetitions (*R_adj_^2^* = .022, *p* = .043). The extent to which current symptoms aligned with the Neurological factor (*ή_p_^2^* = .028) predicted the number of WCST perseveration errors (*R_adj_^2^* = .021, *p* = .047). Alignment with the Cardio-Pulmonary factor (ή_p_^2^ = .035) predicted WCST reaction time of correct answers (*R_adj_^2^* = .028 *p* = .025). None of the factors were associated with any variables within the Associative Memory or Word List tests. After correcting for multiple comparison (Sidak *α* = .0028), no associations were significant.

### 3.7 Cognitive Symptoms and Cognitive Performance

As reported in our previous publication with this sample (Guo et al. 2021), cognitive symptoms were highly prevalent. Within those currently experiencing symptoms (*n* = 126), 77.8% reported difficulty concentrating, 69% reported brain fog, 67.5% reported forgetfulness, 59.5% reported tip-of-the-tongue (ToT) problems and 43.7% reported semantic disfluency (saying or typing the wrong word).

A cognitive symptoms factor was created separately to the noncognitive symptoms (see Guo et al. 2021) for both ongoing and current symptoms. There was no association between the ongoing cognitive symptoms factor and any cognitive factor. In terms of individual cognitive task variables, the ongoing cognitive symptom factor significantly predicted variance in the Word List Recognition Memory Test, with more severe reported cognitive symptoms associated with lower % correct (*ή_p_^2^* = .038; *R_adj_^2^* = .031, *p* = .02) and slower reaction times (*ή_p_^2^* = .039; *R_adj_^2^* = .032, *p* = .018). Ongoing cognitive symptoms were also associated with number of repetitions in the Category Fluency Test (*ή_p_^2^* = .032; *R_adj_^2^* = .025, *p* = .032) and reaction time in the 2D Mental Rotation (*ή_p_^2^* = .029; *R_adj_^2^* = .022, *p* = .042). However, none of these associations survived correction for multiple comparisons (Sidak *α* = .0028).

Current cognitive symptoms significantly predicted variance in the Memory factor (*ή_p_^2^* = .046; *R_adj_^2^* = .039, *p* = .012) only. In terms of individual variables, current cognitive symptoms significantly predicted variance in Word List performance (but not RT) metrics (d’: *ή_p_^2^* = .03; *R_adj_^2^* = .024, *p* = .036; % correct: *ή_p_^2^* = .06; *R_adj_^2^* = .053, *p* = .003), Category Fluency repetitions (*ή_p_^2^* = .048; *R_adj_^2^* = .041, *p* = .009), and 2D Mental Rotation reaction time (*ή_p_^2^* = .041; *R_adj_^2^* = .034, *p* = .015). However, none of these associations survived correction for multiple comparisons (Sidak *α* = .0028).

Some specific cognitive symptoms can be related directly to tests of the associated ability. Participants that reported currently experiencing forgetfulness were compared to those not reporting this symptom on measures of memory. Forgetfulness was associated with a reduced score on the overall memory factor (*t*(134) = 2.111, *p* = .037; Figure 5) even when demographic variables were accounted for (*F*(1,120) = 8.840, *p* = .030). For individual memory variables, those reporting forgetfulness scored significantly lower on the Word List Recognition Memory Test (*U* = 2.48, *p* = .013) but no difference was found for Associative Memory. After controlling for age, sex, country and education level, no differences were significant among the individual variables.

**Figure 5.**
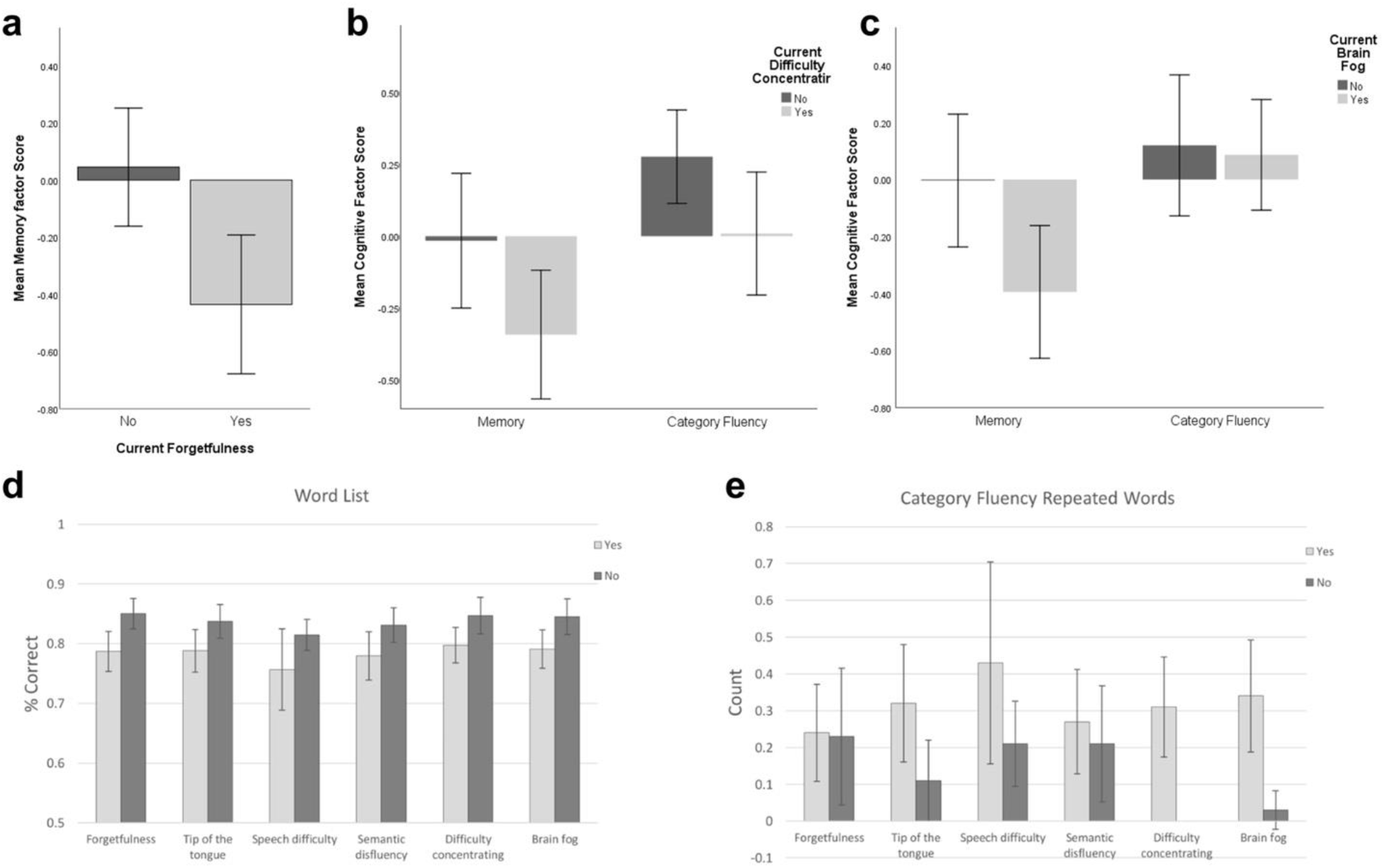
(a) Memory factor score for those reporting (or not) current forgetfulness. (b) Memory and Category Fluency factor scores for those reporting (or not) current difficulty concentrating or (c) brain fog. (d) Word List % correct among groups with/without current cognitive symptoms; e) Category Fluency repetitions among groups with/without current cognitive symptoms. Error bars: +/− 2 SE.

Participants reporting linguistic problems (two cognitive symptoms: ToT, semantic disfluency; one neurological symptom: speech difficulty, e.g., slurring) were compared to those not reporting these symptoms on measures of involving verbal/linguistic challenge. For the Category Fluency factor, there was no effect of ToT (*t*(135) = .414, *p* = .680), semantic disfluency (*t*(135) = .671, *p* = .503) or speech difficulty (*t*(16.4) = .039, *p* = .969). In terms of individual linguistic (Word list and Category Fluency) variables, those reporting ToT problems trended towards lower % correct of Word List Recognition (*U* = 1.91, *P* = .057) and repeated significantly more words on Category Fluency (*U* = 2.22, *p* = .026) than those without this symptom. Those reporting semantic disfluency had significant lower % correct (*U* = 2.49, *p* = .013) and d’ (*U* = 1.99, *p* = .047) on Word List Recognition than those without this symptom. Finally, those reporting speech difficulty had significantly lower % correct on Word List Recognition (*U* = 2.15, *p* = .031) and more repetitions on Category Fluency (*U* = 2.37, *p* = .018) than those not reporting this symptom. Again, after controlling for age, sex, country and education level, no differences were significant.

Finally, to establish whether any cognitive performance differences were due to “general” issues with cognition, we compared individuals experiencing “general” cognitive issues (difficulty concentrating and brain fog) to those not reporting these symptoms across all cognitive tests.

Difficulty concentrating was not associated with variance in any cognitive factor. However, controlling for demographic variables revealed an association between reporting difficulty concentrating and lower scores on the Category Fluency factor (*F*(1,121) = 4.199, *p* = .043). Brain fog was associated with significantly reduced performance on the Memory factor only (*t*(134) = 2.151, *p* = .033) which dropped below significance (*p* = .054) when demographic variables were accounted for. Neither Executive Function factors showed any significant association with these symptoms.

In terms of individual variables, those reporting difficulty concentrating had fewer correct words (*U* = 2.11, *p* = .034) and more repetitions (*U* = 2.74, *p* = .006) on Category Fluency, but had faster reaction time on 2D Mental Rotation (*U* = 2.26, *p* = .024) than those not reporting this symptom. After controlling for age, sex, country and education level, these differences were maintained, with those reporting difficulty concentrating producing fewer correct words (*F*(1,106) = 8.19, *p* = .005), and more repetitions (*F*(1,106) = 4.28, *p* = .04) on Category Fluency, and faster reacting faster on the 2D Mental Rotation Test (*F*(1,107) = 5.68, *p* = .019). However, none of these survived correction for multiple comparisons (Sidak *α* = .0028).

Those reporting brain fog had lower performance on Word List Recognition (*U* = 2.35, *p* = .019) and produced more repetitions in Category Fluency (*U* = 3.04, *p* = .002) than those not reporting this symptom. After controlling for age, sex, country and education level, the difference on Word List Recognition disappeared but those reporting brain fog still had more repetitions on Category Fluency (*F*(1,106) = 6.90, *p* = .01). However, this did not survive correction for multiple comparisons (Sidak *α* = .0028).

## 4 Discussion

Here we present that second subset of initial findings from a cross-sectional/longitudinal study investigating cognition post-COVID-19: The COVID and Cognition Study (COVCOG). In this investigation, we explore how factors associated with COVID-19 infection may impact performance on cognitive tests.

Participants were assessed on a range of cognitive tasks intended to cover different aspects of memory (verbal memory and associative memory), language (word finding), and executive functions (task switching and visual working memory). Our first hypothesis was that those who had suffered COVID-19 infection would be likely to show deficits in tasks challenging memory and language, given the prevalence of self-reported cognitive symptoms in these areas.

We found that the fact of COVID-19 infection (irrespective of ongoing symptoms) was associated with reduced performance on a factor created from memory task variables, but not other cognitive factors (once demographic variables were accounted for). Detailed analysis of individual variables showed an increased reaction time when performing a verbal memory task (alongside a number of other Word List and Associative Memory variables which did not survive correction for multiple comparisons). When considering severity of ongoing symptoms, once again memory emerged as a significant factor, with those with severe ongoing symptoms performing significantly worse than those that had recovered. Looking at individual variables, the impact on verbal memory specifically became clear, with both performance (% correct) and reaction time being significantly affected by severity of ongoing illness in a dose-dependent manner (those with severe symptoms were worse than those with mild symptoms who were worse than those that had recovered). The picture was less clear for non-verbal associative memory, which did not show a main effect (after correcting for multiple comparisons) but in pairwise analyses did demonstrate a clear performance advantage in those who had not suffered COVID-19 infection relative to those with severe ongoing symptoms. The Category Fluency word finding task showed a similar pattern, with main effects falling below the threshold for significant once multiple comparisons were accounted for, but pairwise analysis revealing a strong negative impact of severe ongoing illness on ability to produce category words. Looking to executive functions, similar to Hampshire and colleagues (2021) we found little to no effect of COVID-19 infection on 2D Mental Rotation, which is thought to assess visuospatial working memory (Hyun and Luck 2007). While some group differences emerged in reaction times during the Wisconsin Card Sorting Test, these disappeared after controlling for demographic factors, suggesting they may have been an artefact of the slightly older age of those with ongoing COVID-19 symptoms.

Long COVID is often reported to be a cyclical illness, with symptoms changing in severity over time. As such, it was important to establish whether severity of symptoms on the day of test (rather than in general) might account for significant variance in cognitive performance. We found that the extent to which participants reported that they were having a “bad day” in terms of symptoms on the day of test was not directionally associated with performance on any task, and furthermore did not contribute to models predicting cognitive performance from severity of ongoing symptoms. This suggests that it was the general severity of the ongoing illness—rather than feeling ill on that day in particular—that was driving alterations in cognitive performance.

Given these findings, we suggest that, as others have found (e.g., Hampshire et al. 2021), “objective” cognitive differences do exist between those that have and have not suffered COVID-19 infection. In particular, we find that these are related to the severity of ongoing illness (with those who report having fully recovered being, in our sample, indistinguishable from those who have not had the infection), and that they may be most pronounced in tests of verbal memory. Particular difficulties with language and verbal memory align with frequency of self-reported deficits in these areas in other studies of Long COVID (e.g., Davis et al. 2021; Ziauddeen et al. 2021) as well as evidence for concentration of gray matter loss in the left hemisphere (Douaud et al. 2021).

In our previous publication on the COVCOG sample (Guo et al. 2021), we reported that differences in long-term severity of Long COVID symptoms could be partially predicted by severity and nature of the initial illness. Here we found that reported severity of initial illness did not influence later performance on cognitive tasks taken. However, there was an influence of the *nature* of the initial illness. Using the symptom factors we introduced previously (Guo et al. 2021), we found that individual differences in the initial Dermatological symptom factor predicted around 8% of variance in Executive Functions Reaction Times, while around 5% of variance in Memory was predicted by individual differences in the Fatigue/Systemic initial symptoms factor. These results were reflected in the individual cognitive variables, where the Fatigue/Systemic symptom factor predicted multiple memory variables (e.g., word list d’, % correct and reaction time), while the Dermatological factor predicted Associative Memory and 2D Mental Rotation reaction time. Interestingly, the initial symptom factors predicting cognitive performance were not quite that same as those that were found to predict cognitive symptoms in our previous analysis. There we showed that a model containing all factors *except* the Dermatological symptom factor predicted around 20% of variance in ongoing cognitive symptoms, and that a similar model (omitting Respiratory symptoms) predicted around 14% of variance in current cognitive symptoms. One explanation for the differential findings here may be that measures of reaction-time may not align so closely to individuals’ perceived cognitive issues.

One hypothesis was that neurological symptoms during the acute phase may signal increased likelihood of subsequent cognitive issues. While we found no clear association between the initial Neurological factor and cognitive function, one specific symptom—disorientation— experienced during this period predicted variance in both executive functions and memory. There were also a number of associations between neurological symptoms experienced in the first 3 weeks and individual cognitive task variables (notably headache, altered consciousness and numbness), however these did not survive correction for multiple comparisons. As discussed in our previous report (Guo et al. 2021), the Fatigue/Systemic factor, while not labelled “Neurological” contains a large number of neurological symptoms, including confusion, numbness, headache and dizziness, the latter two of which loaded more highly on the Fatigue/Systemic factor than on the Neurological factor (which was more characterized by disorientation, visual disturbances, delirium and altered consciousness). The fact that it was this factor, rather than the Neurological factor, that predicted later cognitive task performance may be informative as to the mechanism of action. The Fatigue/Systemic factor might be considered to incorporate many of the expected features of systemic inflammation, in contrast to the Neurological factor that is more closely linked to neurological system only. This account accords with the other factors that emerged as predictors. While named for the fact that they affect the skin, the symptoms in the “Dermatological” factor are also linked with systemic inflammation, incorporating cross-loading symptoms such as limb weakness. These findings suggested that systemic inflammation associated with acute COVID-19 infection may have contributed to cognitive deficits across different domains up to 6 months later.

In terms of ongoing symptoms, the main finding to emerge was that the Neurological factor predicted variance in Executive Function Performance, perhaps driven by an influence of this cluster of symptoms on the Wisconsin Card Sorting Test (though no individual task variable survived correction for multiple comparison). The Neurological factor also emerged as significant predictor of cognitive performance among the current symptoms, this time significantly predicting variance in Memory. These associations align to some degree with the previous finding that current cognitive symptoms were well predicted by models containing ongoing Neurological, Gastrointestinal and Cardio-Pulmonary symptoms, and current Neurological and Cardio-Pulmonary symptoms (Guo et al. 2021). The shift in predictive power from predominantly inflammatory variables during the acute phase, to more classic neurological symptoms during the ongoing illness raises the possibility that damage or processes instigated by an excessive immune response to infection may lead to disruption of neural function with neurological and cognitive consequences that linger independently. Such a mechanistic hypothesis would require targeted investigation of inflammatory markers, as well as functional and structural imaging.

As has been noted, the symptom factors that predicted performance on cognitive tasks were not always the same as those that predicted individual differences in cognitive symptoms. Indeed, individual differences in ongoing cognitive symptoms did not predict variance in any cognitive task performance factor. Currently experienced cognitive symptoms were associated with reduced memory performance, driven by differences in multiple verbal memory tasks (particularly Word List and repetitions within the Category Fluency Test). When investigating specific cognitive symptoms, those who reported currently experiencing forgetfulness showed significantly lower Memory factor score, while those reporting linguistic issues did not score differently on the Category Fluency factor (although there were some associations with individual Category Fluency and Word List task variables that did not withstand controlling for demographic factors). The finding that those currently reporting cognitive issues—particularly memory problems—scored significantly lower on objective cognitive tasks than those experiencing ongoing symptoms but *not* reporting such symptoms, and that both are linked with ongoing neurological symptoms is important. It suggests that subjective experience of cognitive deficits in this population may be considered predictive of need of neurological assessment and treatment.

Many of the limitations of this study have been reviewed in our previous report (Guo et al. 2021). One major limitation of this study is that due to the novelty of the topic, it was not designed with clear, specific hypotheses, and as such much of the analysis was necessarily exploratory, resulting in a large number of analyses and comparisons. To account for these, Sidak alpha adjustments were used, with the result that only the very strongest effects survived at conventional statistical thresholds. We consider this conservative approach appropriate, but note that it is likely to be associated with a high type 2 error rate—and thus that some associations that did not reach these thresholds may yet be upheld upon further investigation/replication. A stated aim of this study was to generate hypotheses that could be tested in later, more targeted research, and thus while only the strongest statistical outputs should be treated as concrete findings, those that do not reach this threshold are also reported, such that they can inform and motivate future research. Of particular note is that, while rarely surviving corrections for multiple comparisons, variables associated with the Word List Recognition Memory Test repeatedly emerged as being modulated by facets of Long COVID. This is particularly relevant since it was predominantly this task that was influenced by severity of ongoing symptoms. All elements of this task (performance and reaction time) were predicted by Fatigue/Systemic symptoms during the initial illness, and performance was related to ongoing Cardio-Pulmonary symptoms, and current Neurological symptoms and Appetite Loss. Word List performance was also linked with severity of cognitive symptoms both ongoing and current. The consistent implication of verbal memory as vulnerable to factors associated with COVID-19 infection should certainly warrant further, more targeted investigation.

Another potentially notable finding that may be somewhat obscured by alpha corrections is the consistency in the association between neurological symptoms and executive function, particularly within the Wisconsin Card Sorting Test. While the more “encephalitis-like” Neurological initial symptoms factor did not show associations with later WCST performance, individual elements of it (dizziness, disorientation, numbness) did. As already stated, during the ongoing illness, the neurological factor strongly predicted number of perseveration errors, but was also associated with reduced correct responses and slower reaction times. This pattern was carried over into currently experienced symptoms, with neurological symptoms once again predicting perseveration errors. Taken as a pattern, these findings (though not all individually strong) may suggest that more severe neurological symptoms may be indicative of alterations in frontal lobe function, evidenced by problems with response inhibition. This, again, should be investigated in more targeted future studies.

An additional limitation of this study was that data was collected online. While online assessment facilitated cognitive testing during lockdown—and with patients from around the world—it meant that we were less able to guarantee high quality data by ensuring that participants were in a suitable environment or concentrating properly on the task. This was mitigated to some degree by the use of the “concentration/bot check” task which did not highlight a problem with lack of concentration. It is also increasingly become accepted that online cognitive testing can produce highly robust and reliably results, and that Gorilla.sc is a reliable platform on which to conduct this type of research (e.g., Anwyl-Irvine et al. 2020; Hilbig 2016). Nonetheless, future research should confirm these findings using full lab-based cognitive testing batteries.

Our study contained very few individuals who fell at either end of the severity spectrum (e.g., were asymptomatic or required ventilation). The deficits identified in Hampshire and colleagues’ (2021) study were substantial and related to severity, with ventilated participants showing performance reductions larger than those seen (using the same tasks) following a stroke, and greater than the average 10-year decline. They also found that detectable deficits were also present in those that experienced no respiratory symptoms at all, and did not have ongoing symptoms. In contrast, our present results suggest that those who report being completely recovered from COVID-19 were indistinguishable from those that had not suffered infection at all. This difference may be due to the relative power of the two studies (with Hampshire and colleagues having a large sample). It may also be related to differences in how symptomatology was recorded. Hampshire and colleagues only asked about “breathing difficulties” in the initial illness, and their assessment of ongoing symptoms was a sub-choice within “have you had, or suspect you have had symptoms of COVID-19?” (“No” / “Yes but the symptoms passed” / “Yes currently experiencing symptoms”). Given that people’s experience of symptoms during the long-term sequalae of COVID-19 can be very different to the “Classic” COVID-19 symptoms of breathing difficulties, cough and loss of sense of taste and smell, many individuals who were experiencing, for example, ongoing cognitive or neurological symptoms may not have considered these to qualify in this context. Further research will be necessary to clarify these discrepancies.

### 4.1 Long term risks

The accumulating neural and cognitive findings in Long COVID patient groups present a concerning picture when considering long-term cognitive health. In particular, loss of gray matter within the temporal lobe in COVID-19 (Douaud et al. 2021), along with the evidence for reduced memory performance presented here, supports the suggestion that those who have suffered COVID-19 infection may be at increased risk for later neurodegeneration and dementia (de Erausquin et al. 2021).

While some authors have particularly highlighted the neurodegenerative risks posed via viral invasion of the central nervous system (CNS) (Douaud et al. 2021), in fact almost all candidate mechanisms of neural impact raise the possibility of increased vulnerability to dementia. SARS-CoV-2 is increasingly being recognised as an inflammatory disease (Pearce, Davidson, and Yellon 2020; Sims et al. 2021). As well as having major physical impacts, excessive and chronic inflammation is also associated with considerable damage in the brain. Chronic neuroinflammation is heavily implicated in the pathophysiology of neurodegenerative diseases (Chen, Zhang, and Huang 2016), with evidence of inflammation commonly being found in the brains of patients suffering from Alzheimer’s disease (AD) (McGeer and McGeer 2010; Zotova et al. 2010). The dramatic impact of infections such as E-coli on survival and proliferation of hippocampal neurons (Ekdahl et al. 2003; Monje, Toda, and Palmer 2003) has previously indicated that this region may be vulnerable to deleterious effects of inflammatory viral infection, and development of dementia following viral infections such as influenza have been previously noted (e.g., Honjo, van Reekum, and Verhoeff 2009). COVID-19 has also been linked to abnormal blood clotting, which again has been linked to disease severity and death (Tang et al. 2020; Wang et al. 2020; Wichmann et al. 2020; Xiang-Hua et al. 2010), with microthrombi in multiple organs, including the brain (Zhang, Tecson, and McCullough 2020). Clotting is a significant factor when considering risk for neurological damage and cognitive impairment because of the risk of CVAs and stroke (e.g., Klok et al. 2020). Indeed, increased incidence of stroke has been reported in hospitalized COVID-19 patients (Li et al. 2020; Oxley et al. 2020). A large proportion of stroke survivors suffer cognitive impairment, and, unlike physical impairments, these tend to worsen rather than improve over time, leading to the description of “post-stroke dementia” (Mijajlović et al. 2017). Many small stoke events (“transient ischemic attacks” (TIAs)) go unnoticed at the time but may cause cumulative damage leading to cognitive decline and dementia vulnerability. Indeed, recent studies have indicated that the proportion of dementia that is caused by small vessel ischemia may be as high as 36–67% (e.g., Grau-Olivares and Arboix 2009; Seshadri and Wolf 2007).

### 4.2 Summary

In this second investigation of the first baseline session of the COVID and Cognition study, we explored whether those who had suffered COVID-19 infection showed measurable differences in assessments of cognitive performance. We found a consistent association between COVID-19 infection and reduced memory performance, with those with ongoing symptoms were less accurate and slower in a test of verbal memory, but (once demographics and multiple comparisons were accounted for) there were no significant group effects in any other cognitive domain. When considering the nature of symptoms experienced, Fatigue/Systemic and Dermatological symptoms during the initial 3 weeks of illness were associated with reduced memory performance and slower reaction times on executive function tasks respectively. Neurological symptoms during the ongoing illness were associated with performance in the executive function task, while the same symptoms experienced at the time of test predicted variance in memory. These were the most robust findings, with a conservative correction for multiple comparisons, suggesting that other identified associations may be worthy of further investigation.

In combination with previous evidence for cognitive dysfunction (e.g., Hampshire et al. 2021) and neural damage following COVID-19 infection (Douaud et al. 2021), these findings are concerning and suggest that COVID-19 is an illness that may be associated with considerable cognitive and neurological sequalae of unknown longevity. This is particularly concerning given the potential for these changes to translate into greater vulnerability to neurodegeneration. The COVID and Cognition participants were followed up multiple times following this assessment, and future publications with this cohort will prove informative as to the likely progression in symptoms and cognitive performance over time. However, given the associations shown in our previous publication with number of weeks since infection (Guo et al. 2021), it seems likely that a considerable proportion of individuals may show stable cognitive symptoms over many months.

## Data Availability

All data produced in the present study are available upon reasonable request to the authors and will be published online soon.

## Acknowledgements

The COVID and Cognition study has benefitted from help and support from a large number of individuals, not least the participants who gave their time. We would especially like to thank members of the Long COVID support group, particularly Claire Hastie, Barbara Melville-Johannesson and Talya Varga for your time and insights. Thanks also to Emma Weisblatt, Mirjana Bozic and Keir Shiels who gave valuable suggestions, to Honor Thompson, Aashna Malik, Connor Doyle, Abel Ashby, Seraphina Zhang for research support and to Helen Spencer for statistical assistance. This study was not supported by any funding bodies but did benefit from research funds from the University of Cambridge department of Psychology.

