## Supplementary Materials for "COVCOG 2: Cognitive and Memory Deficits in Long COVID: A Second Publication from the COVID and Cognition Study"

**Supplementary Table 1.** Rotated component matrix for cognitive task variables

| **Rotated component matrix[a]** | **Component** | |
|---|---|---|
| | **EF** | **RT** |
| **WCST_Correct** | 0.941 | -0.103 |
| **WCST_Persev_error** | -0.888 | 0.162 |
| **WCST_Nonpersev_error** | -0.627 | -0.131 |
| **WCST_RT_correct** | -0.278 | 0.807 |
| **WCST_RT_Nonpersev** | 0.032 | 0.563 |
| **@2D_CorrectPercentage** | 0.585 | 0.111 |
| **@2D_RT_overall** | 0.252 | 0.734 |
| | **Memory** | **Category Fluency** |
| **WL_CorrectPercentage** | 0.84 | 0.116 |
| **WL_RT_overall** | -0.501 | 0.194 |
| **Category_correct** | 0.484 | 0.446 |
| **Cetagory_related** | 0.172 | -0.722 |
| **Category_incorrect** | -0.025 | -0.661 |

| | | |
|---|---|---|
| **Category_CorrectPercentage** | 0.114 | 0.885 |
| **AM_correctPercentage** | 0.475 | 0.216 |
| **AM_RT_overall** | -0.445 | 0.192 |
| **WL_d** | 0.816 | 0.069 |

Extraction Method: Principal Component Analysis.

Rotation Method: Varimax with Kaiser Normalization.

[a] Rotation converged in 3 iterations.

**Supplementary Table 2.** Group and pairwise comparisons including both No COVID and severity levels for cognitive factors.

| **Comparing No COVID, Recovered, Ongoing (Mild/Moderate) and Ongoing (Severe) groups** | | | | |
|---|---|---|---|---|
| | **No COVID** | **Recovered** | **Mild/Moderate** | **Severe** |
| **Factor 1: EF performance** | 1.318 (3,317) | .268 | 0.524 (3,303) | .666 |
| **Factor 2: EF Reaction Time** | 3.719 (3,307) | .012* | 1.422 (3,293) | .236 |
| **Factor 3: Memory** | 6.688 (3,304) | <.001*** | 8.453 (3,290) | <.001*** |
| **Factor 4: Category Fluency** | 2.707 (3,313) | .045* | 0.982 (3,288) | .402 |
| **Pairwise Comparisons** | | | | |
| **Factor 3: Memory** | **No COVID** | **Recovered** | **Mild/Moderate** | **Severe** |
| **No COVID** | - | ns | $t(87.6) = 2.4$, $p = .018$ | $t(99.8) = 3.9$, $p < .001$ |
| **Recovered** | - | - | ns | $t(99) = 2.885$, $p = .005$ |
| **Mild/Moderate** | - | - | - | ns |
| **Severe** | - | - | - | - |
| **Factor 4: Category Fluency** | **No COVID** | **Recovered** | **Mild/Moderate** | **Severe** |
| **No COVID** | - | ns | ns | $F(1,152) = 3.051$, $p = .003$ |
| **Recovered** | - | - | ns | ns |
| **Mild/Moderate** | - | - | ns | ns |
| **Severe** | - | - | - | - |
| **Pairwise Comparisons Controlling for Age, Sex, Country and Education** | | | | |

| **Factor 3: Memory** | **No COVID** | **Recovered** | **Mild/Moderate** | **Severe** |
|---|---|---|---|---|
| **No COVID** | - | ns | $F(1,191) = 5.515$, $p = .02$ | $F(1,205) = 20.38$, $p < .001$ |
| **Recovered** | - | - | ns | $F(1,85) = 6.65$, $p = .012$ |
| **Mild/Moderate** | - | - | - | ns |
| **Severe** | - | - | - | - |

**Supplementary Table 3.** Initial phase neurological symptoms and subsequent cognitive performance.

| **Symptom Factor (Predictor)** | **Cognitive Outcome (dependent variable)** | ***F*** | ***p*** | ***β*** | **Adjusted $R^2$** |
|---|---|---|---|---|---|
| **Headache** | Word List (RT) | (1,158) = 7.92 | .005 | -.219 | .042 |
| | Associative Memory (% Correct) | (1,159) = 4.48 | .036 | .165 | .021 |
| | Category Fluency (Correct) | (1,156) = 8.87 | .003 | .0232 | .048 |
| **Confusion** | Category Fluency (% Correct) | (1,155) = 4.00 | .047 | .159 | .019 |
| | Word List (% Correct) | (1,158) = 7.83 | .006 | .217 | .047 |
| **Altered Consciousness** | Word List (d') | (1,158) = 9.14 | .003 | .234 | .049 |
| **Dizziness** | WCST (persev. errors) | (1,156) = 4.54 | .035 | -.168 | .028 |
| **Disorientation** | WCST (Correct) | (1,156) = 5.57 | .019 | .186 | .034 |
| **Numbness** | WCST (RT Correct) | (1,156) = 5.57 | .003 | -.234 | .055 |

* denotes *p* values below Sidak-correct alpha at .0028
